# The contribution of population age-sex structure to the excess mortality estimates of 2020–2021 in Denmark, Finland, Iceland, Norway, and Sweden

**DOI:** 10.1101/2022.11.18.22282495

**Authors:** Kasper P. Kepp, Jonas Björk, Louise Emilsson, Tea Lallukka

## Abstract

**Background:** The Nordic countries are an ideal case study of the COVID-19 pandemic due to their comparability, high data quality, and variable responses. Excess mortality is a key metric but it is sensitive to data quality, model assumptions, and population structure, with diverse estimates published so far.

**Methods:** We investigated the age- and sex-specific mortality patterns during 2020−2021 for the five Nordic countries using annualized age- and sex specific death rates and populations. We compared the total age- and sex-adjusted excess deaths, ratios of actual vs. expected death rates, and age-standardized excess death estimates. We estimated excess deaths with several time periods and sensitivity tests, using 42 sex and age groups. Our models are less sensitive to outlier years than models based on 5 years of data.

**Results:** Age-specific death rates have declining trends that reflect real improving health demographics. Our total excess mortality is close to WHO’s estimates, except higher for Norway and lower for Sweden, partly due to data used. Total excess deaths were dominated by the age group 70−89 years, was not identified in children, and more pronounced in men than women. Sweden had more excess deaths in 2020 than 2021 whereas Finland, Norway, and Denmark had the opposite. Denmark has the highest death rates before and during the pandemic, whereas Sweden in 2020 had the largest mortality increase. The age-standardized mortality of Denmark, Iceland and Norway was lowest in 2020, and 2021 was one of the lowest mortality years for all Nordic countries. We show that neutral baseline methods underestimate excess deaths and we document the importance of outlier mortality years.

**Conclusions:** We provide excess mortality estimates mortality of the Nordic countries in relation to sex and age, with several metrics important in combination for a full understanding and comparison of the countries. We additionally identify important effects such as mortality displacement and sensitivities that affect our estimates and those of other excess mortality models.

## Introduction

All-cause excess death estimates (total observed deaths minus deaths expected for a given period) include all deaths, and therefore they are not affected by testing and COVID-19-death reporting strategies between countries or territories.^1–5^ Thus, they offer a less confounded basis for comparing total mortality outcome of a health crisis such as the pandemic caused by the severe acute respiratory syndrome coronavirus 2 (SARS-CoV-2) to the deaths expected without a crisis. However, even if total deaths are often known, determining the expected deaths (the baseline in absence of crisis) involves considerable uncertainty.^6–9^

Recently, estimates of the excess mortality of the pandemic years 2020 and 2021 have been published by the World Health Organization (WHO)^10,11^ and the Institute for Health Metrics and Evaluation (IHME).^12^ The Economist^13^ and World Mortality Dataset (WMD)^14^ have continuously provided updated excess mortality estimates. We recently analyzed and compared these models and reported major sensitivity to time-period used due to outlier mortality years and concern about the IHME model.^6^ We also urged strong caution in interpreting policies from the complex and heterogeneous results, and because these estimates should consider population structure, including the changes in the sizes of population age groups over time.

Expected deaths, and therefore excess deaths, are modeled based on expectations that can include covariates such as age or sex. Mortality is most strongly related to age,^15^ and thus changes in the size of the age subgroups over time (especially the oldest most mortal groups) affect baselines and excess mortality estimates.^16^ Annual age-specific death rates (deaths divided by mean populations in a population group) can be used to establish excess mortality in a context of age structure. Instead, the total excess death loss itself is not meaningfully compared between regions or population groups. In the same way, sex-specific mortality may affect total mortality in an age-dependent way that can be accounted for by comparing age- and sex-specific death rates, as recently attempted in a study for Italy specifically^17^.

The most important studies so far in this direction are the age- and sex specific life-expectation studies^16,18^, which also included the five Nordic countries and confirmed them as having some of the lowest mortality impacts overall in Europe, while also documenting the mortality reversal from 2020 to 2021^16^. Previous age- and sex-adjusted estimates for Sweden specifically for the first COVID-19 wave in 2020 have also been published^19^. These studies, as many others, used 5-year pre-pandemic data for building baselines.

In this paper, we aimed to explore the impact of age-sex structure of the population on excess mortality for the five Nordic countries during 2020 and 2021. We studied the Nordic countries as an ideal focused test case due to 1) their unique combination of commensurable and complete health register data, to rule out data weaknesses; 2) high epidemiological comparability that reduces confounding effects on mortality that could add noise to broader country comparisons.^6,20,21^ The Nordic countries are also of interest for analyzing pandemic response due to heterogeneity in response on this data background.

## Methods

### Data

All data used were register-based, collected from the administrative records of the five Nordic countries (see Data availability section), and curated via the Nordic Council’s data page. The data from individual departments are the same as in this source.

The age- and sex-specific death rates are given as deaths in age and sex group in the year divided by the mean population of the age and sex group in the same year, collected for total population, and men (M) and women (F) separately.

These data were collected for 2010−2021 (12 years). Years 2010−2019, 2010−2018, and 2015−2019 were used for extrapolating trends. Excess deaths were calculated for 2020 and 2021 separately based on extrapolations from the data until 2019 and 2018. Mean populations and death rates in 1-year groups were combined into 5-year groups, except 0−5 years, which was divided into <1 years and 1−5 years, due to the special high mortality of newborns, which makes the <1 year mortality group very distinct from children 1−4 years. Similarly, due to low population size and the associated statistical noise, age groups above 95 years were combined, to a total of 21 age groups for men and women, i.e., 42 subgroups per country.

We used the final annualized deaths, mean populations, and death rates within each age and sex group to avoid issues with e.g., ISO week (International Organization for Standardization), which can affect excess death estimates by having years of different length. When extrapolating based on full years, 2015 (starting point for many interpolations) and 2020 were ISO-leap years with 53 ISO-weeks with corresponding additional deaths, whereas the years 2016−2019 had 52 ISO weeks and correspondingly fewer deaths in the ISO calendar (**Table S1**).

### Estimating expected deaths

The excess deaths *D*_ex_ for the population subgroup *i* are defined as:

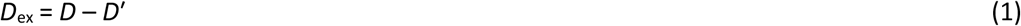

Where *D* are the actual deaths observed in the time-period for the population subgroup of interest, and *D*’ are the expected deaths of the same population group during the same period. *D* can also be written as:

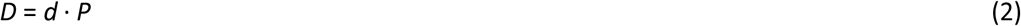

*P* is the mean size of the population sub-group *i* during the time-period, and *d* is the death rate for the group. To account for the impacts of population, the expected deaths *D*’ also need to be decomposed into population subgroups via an expected death rate *d*’ and an expected population size in absence of crisis, *P*:

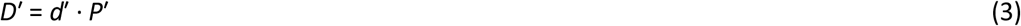

Since the size of the population subgroup is a function of the death rate, for old groups with high mortality and small populations, one can expect an error if using the observed mean populations *P*, and thus it is of interest to establish the error (referred to as the *population error*) made if assuming *P*’ = *P*:

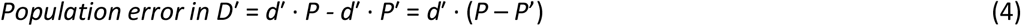

In a crisis with substantial excess mortality, since *P* < *P*’, the population error can lead to underestimation of expected deaths and overestimation of excess deaths. To test the impact of population changes during the crisis period, *P*’ was applied both as the mean population of each subgroup in 2020 and 2021, respectively, when computing the 2020 and 2021 excess deaths, and as the start populations January 1, of 2020 and 2021. It should also be noted that the appropriate choice of P’ may depend on whether the full crisis period or perhaps a part of the period is of interest.

Linear regression of the trends in age- and sex-specific death rates for 2010−2019 were used to estimate expected death rate in each age group for 2020 and 2021. The linear method avoids oversensitivity to gradient-based splines and includes some of the population structure effects (thus having the least change when doing age-correction)^8^ and averages out mortality displacement^22–24^ that seems important to the Nordic countries^25^ by producing a compromise amid the fluctuations.

Also, the years 2018 and 2019 were unusual mortality years for some Nordic countries, e.g., Sweden and Denmark, and 5-year trends are more sensitive to these years and give results distinct from 10-year-extrapolation.^6^ The linear 10-year trends in the age-specific death rates justify the use of 10-year over 5-year trends. Since mortality increases steeply with age, shorter age intervals increase estimate precision. This explains our strategy of using 10-year linear trends specified on sex and 5-year age groups.

### Calculating excess deaths

The excess death count was calculated from the sum of the following terms from all subgroups:

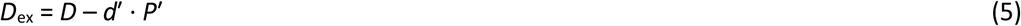

Since population cycles can be noisy (with cyclic and linear components) extrapolating expected population sizes absent a crisis is uncertain. The impact will be largest for the small groups of high age with most mortality, with some methods attempting to correct for this^26^. We calculated *D*_ex_from two different *P*’ (mean population or start population of the crisis year) to investigate impact of population error in (4) and several different *d*’ reflecting inclusion or exclusion of timer periods in the estimates.

The extrapolated expected death rate of each subgroup *d*’ was calculated using average projection, linear extrapolation from 2010−2019, from 2015−2019, and from 2010−2018 (to study the impact of the mortality year 2019 on extrapolations), to establish the uncertainties in the estimates due to variations in the death rate trends affected by pre-pandemic mortality fluctuations. Excess death estimates were obtained using the expected death rate multiplied by the actual mean population size of 2020 and 2021 for each population subgroup, providing an estimate of the excess deaths in persons for the subgroup.

We compared the excess deaths in three ways: 1) Crude excess deaths obtained from death rate extrapolations (age-and sex adjusted total excess deaths), 2) death rates relative to expectations for the countries in their individual historic context since many drivers may contribute to country-specific mortality patterns (age specific mortality ratios), and 3) direct comparison of death rates, standardized to the 2020 Danish population to facilitate country comparisons (age-standardized excess deaths).

The method applies the knowledge of the actual populations of 2020 and 2021 for each specific sex and age population subgroup, but the use of either start or mean populations shows that this choice is not critical (i.e., the population error is small in this case, see results).

## Results

### Trends in Nordic population age structure relevant to excess mortality estimates

The age structure of the Nordic countries differed somewhat at the onset of the pandemic, making age correction important (**Figure S1**). Notably, the 70−74-year age group was as large as younger groups in Finland, somewhat smaller in Denmark and Sweden (plateau-shaped distributions), but considerably smaller in Norway and Iceland with more gradual shifts in age group sizes, and monotonic increase over recent years among 75−79-year-olds that largely contribute to mortality. In 2020, the percentage of the full populations being 70 years or above were 9.8% in Iceland, 12.6% in Norway, and 14.5, 14.9, and 16.1% in Denmark, Sweden, and Finland.

In addition to these effects, periodic birth cycles (variations in the size of birth cohorts^27,28^) and migration effects affect the population structure over the time-period used to estimate expected deaths (**Figures S2−S3**) and can be accounted for by using age-specific death rates for each year specifically taking into account the age group sub-population changing non-monotonically over time.

The trends in age-specific death rates for the age groups dominating total deaths justify linear extrapolation are shown in **Figure 1** (with examples of regression lines shown in **Figure S4**). This also indicates (as discussed in more detail below) that it is the non-monotonous changes in age-specific populations due to birth cycles that produce the more complex age-dependencies on total excess death estimates. Denmark had slightly higher overall death rates for most age groups. Since the age-specific death rates display declining trends, methods using neutral baselines, e.g., pre-pandemic averages without trend^29^ may under-estimate excess deaths (by overestimating expected deaths), as we show in detail below.

**Figure 1.**
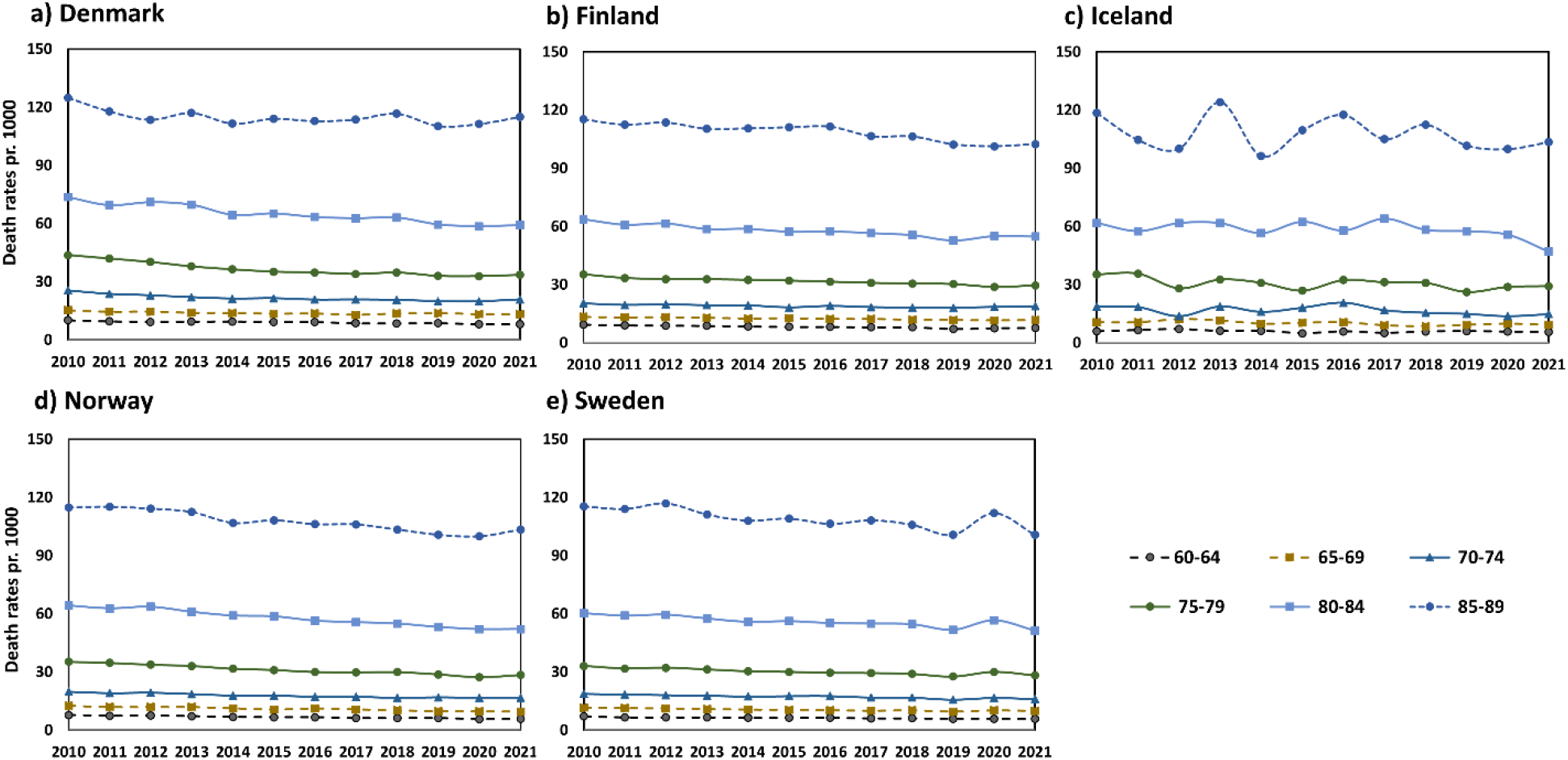
Age-specific death rates per 1000 people for the Nordic countries in 2010−2021 for the age groups mostly contributing to total excess deaths (60−89y). **A)** Denmark. **B)** Finland. **C)** Iceland. **D)** Norway. **E)** Sweden. Declining trends will cause average death rate extrapolation to overestimates expected deaths.

### Estimates of age-specific total excess mortality for the Nordic countries

**Figure 1.** shows the age-specific death rates for the age groups that contribute the most to the deaths, due to their combined mortality and population sizes. Despite the variation in crude death rates, the pre-pandemic trends indicate a tendency of improving health demographics over recent years for the age groups contributing mostly to total deaths. This is observed also for Finland despite its increasing crude death rates, so this is a general Nordic tendency. These trends need to be accounted for when estimating expected deaths in each age group, as done with our methodology.

**Table 1** summarizes the main results from our method. The crude estimates use the crude death rates (without age specification) which account for total mean population changes over time, which is why they differ somewhat from the 10-year estimates published previously that, as other models, used crude deaths without account for changing populations over time. The additional estimates are derived from summing the excess deaths from all the subpopulation groups, either 21 (age-weighted) or 42 (age- and sex-weighted), i.e., accounting for changes in the size of each subpopulation group during the pre-pandemic extrapolation period and during the pandemic. We note that the estimates for Iceland are less certain due to small total numbers.

**Table 1.** Estimated total excess deaths from expected deaths in 2020 and 2021 extrapolated from linear trends in death rates (2010−2019 except * based on 2010−2018) using mean subgroup populations.

|  | Denmark | Finland | Iceland | Norway | Sweden |
| --- | --- | --- | --- | --- | --- |
| Crude |  |  |  |  |  |
| 2020 | 1018 | 772 | -98 | 681 | 7505 |
| 2021 | 3460 | 2557 | -121 | 2411 | 1774 |
| **2020+2021** | **4478** | **3330** | **-219** | **3092** | **9279** |
| Age-weighted |  |  |  |  |  |
| 2019* | 590 | -711 | -57 | 200 | -3462 |
| 2020 | 893 | 597 | -50 | 37 | 7040 |
| 2021 | 3270 | 2299 | -75 | 1421 | 810 |
| **2020+2021** | **4163** | **2896** | **-124** | **1459** | **7850** |
| Age/sex-weighted |  |  |  |  |  |
| 2020 | 937 | 666 | -50 | 79 | 7146 |
| 2021 | 3340 | 2399 | -72 | 1485 | 974 |
| **2020+2021** | **4277** | **3065** | **-122** | **1564** | **8120** |

Norway, Sweden, and Iceland had similar expected death rates for the 75−79-year age group (27.5−27.6 per thousand for 2020 and 26.9−27.0 for 2021, due to the improving health trend, **Figure S4**), whereas for Finland they were 29.5 and 29.1 per thousand, and for Denmark they were 30.8 and 29.7. In other words, death rates tend to be higher in Finland and Denmark than the other countries for this age group. Age correction had a particularly large effect for Norway (**Table 2**). This was mainly due to people 70−79 years old where Norway had estimated 593 excess deaths while Denmark had 2514 for 2020−2021. For the 75−79-year age group, Denmark and Norway have 1512 and 204 excess deaths, respectively. While Norway generally has slightly lower death rates for these age groups, the declining pre-pandemic trend is stronger in Denmark than other countries (**Figure S4a** and **S4d**).

**Table 2.** Total estimated excess deaths per 1 million population (mean population of 2020 and 2021).

|  | Denmark | Finland | Iceland | Norway | Sweden |
| --- | --- | --- | --- | --- | --- |
| Crude |  |  |  |  |  |
| 2020 | 175 | 140 | -268 | 127 | 725 |
| 2021 | 591 | 462 | -325 | 446 | 170 |
| 2020+2021 | **765** | **601** | **-594** | **572** | **895** |
| Age-weighted |  |  |  |  |  |
| 2020 | 153 | 108 | -136 | 7 | 680 |
| 2021 | 558 | 415 | -200 | 263 | 78 |
| 2020+2021 | **711** | **523** | **-336** | **270** | **758** |
| Age-weighted and standardized |  |  |  |  |  |
| 2020 | 152 | 116 | -205 | 9 | 624 |
| 2021 | 546 | 379 | -310 | 287 | 91 |
| 2020+2021 | **698** | **495** | **-515** | **296** | **715** |

Norway and Denmark had the largest relative shifts of population into the 75−79-year age group compared to other countries (**Figure S5**), i.e., their crude excess death rates become relatively smaller once this is accounted for. However, for Denmark this tendency is partly compensated by the steep decline in expected death rates (**Figure S4**), lowering expected death rates more for Denmark, giving higher final excess deaths than for Norway. For Finland and Sweden, the two tendencies (more slowly declining death rates as Norway, **Figure S4**, and somewhat less populations shifted into the mortal age groups in 2020−2021 compared to Norway) make the effect of age correction relatively smaller than for Norway (Iceland’s data also suggest an effect but are uncertain). Thus, age correction had large effect when the population of high-mortality groups changed in a way that was not compensated by trends in death rates.

The numbers in **Table 1** are not associated with confidence intervals since time-period and method variations affect estimates more than the extrapolation uncertainty, which thus underestimates true uncertainties: For relevant uncertainties, we instead refer to the sensitivity tests below. Importantly, these excess crude deaths should be directly seen in the context of the rapidly increasing population sizes of the 75−84-year age groups over recent years (**Figure S2**), and thus, excess death rates, rather than excess deaths, are more appropriate for country comparison, as done below.

To make the crude excess deaths more comparable, the estimates of **Table 1** are listed per million people in **Table 2**. These estimates allow comparison of the total excess death burden per million for the countries but cannot be used to compare the relative success of the countries, which requires expectations based on population age structure, especially regarding older population groups, as done below.

Total excess death estimates, even if age-adjusted, do not allow a comparison of country performance since they hide context of population structure, even if the excess death estimates were accurate, disregarding that very different estimates have been published.^6,10,12–14,29^ We can instead compare death rates directly and make these death rates more relatable by weighting with a standard population such as that proposed by WHO^30^, which in **Table S2** is compared to the Scandinavian standard population by Doll and Cook^31^ and the population of Denmark in 2020.

The real populations deviate substantially from both standard populations, and thus, standard deaths (death rate multiplied by standard population of the age group) do not reflect a true mortality burden. We therefore used the Danish 2020 population as standard population and converted age-standardized mortality rates into crude deaths per million that would have occurred since 2010 if all the other populations had a Danish 2020 population (**Figure 2**). This puts the pandemic excess deaths in a historic comparative context, and confirms the large mortality shifts in Sweden during 2019, 2020, and 2021.

**Figure 2.**
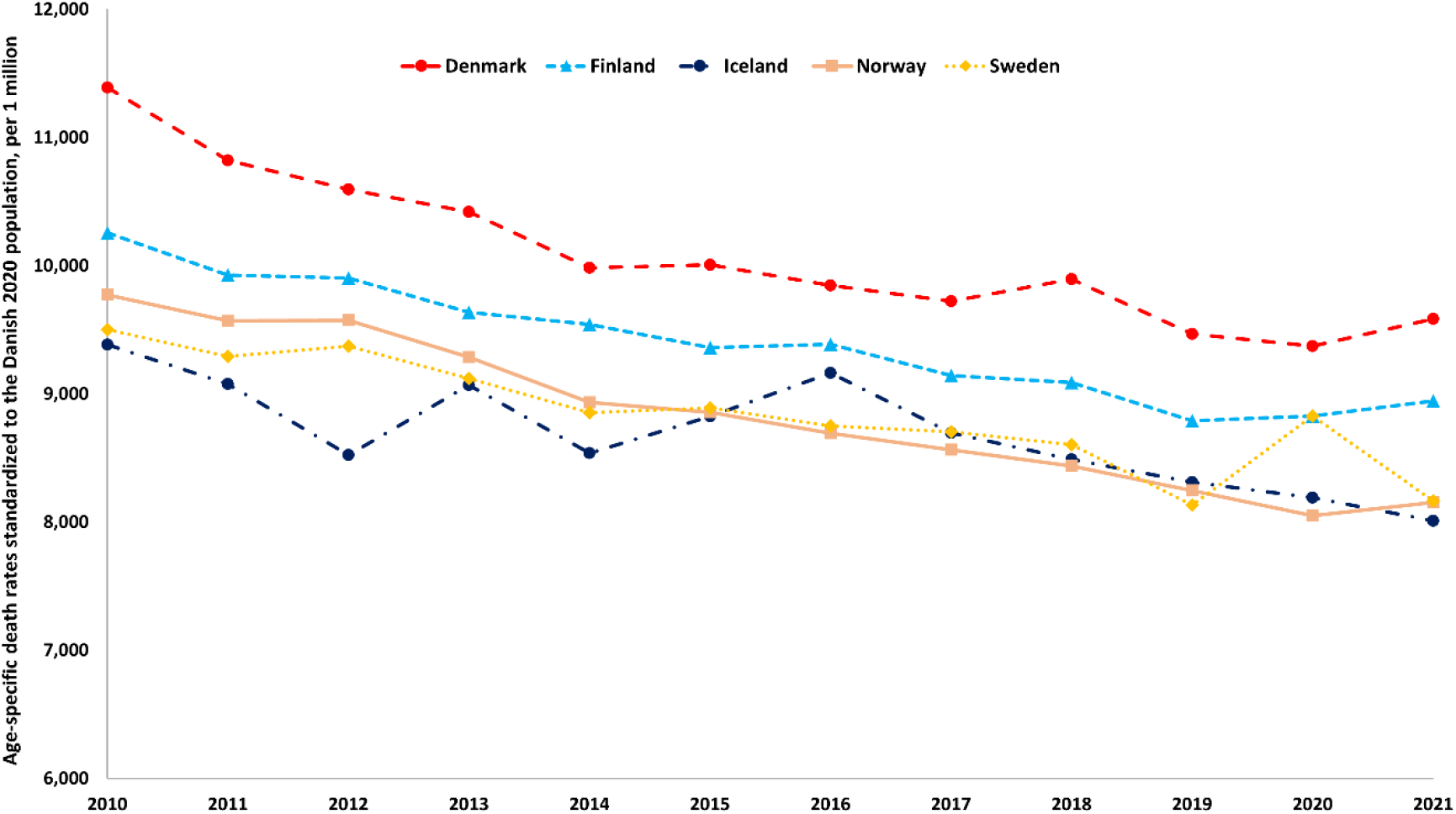
Age-standardized total deaths per million, if all countries had fixed 2020 Danish populations.

Denmark had the highest age-standardized death rates both before and during the pandemic, whereas Sweden briefly reached Finnish death rates in 2020. The age-standardized mortality of Denmark, Iceland and Norway was lowest of all data in 2020, following the historic trend, and despite mortality reversal 2021 was still at least third-best for all Nordic countries. For Sweden, the high mortality in 2020 was comparable to 2015 but lower than the years before 2015. The age-specific and age-standardized excess death estimates derived from the linear 10-year trends (**Figure S6**) are shown in the last part of **Table 2**.

### Sex-specific mortality patterns

We also analyzed sex-and age-weighted mortality, redoing all extrapolations separately and recalculating expected deaths and excess deaths for men and women (**Table 3**). The age- and sex-specific excess death show that for all five countries, men had higher pandemic excess mortality than women, consistent with the picture observed for registered COVID-19 deaths, with most excess deaths arguably being due to Covid-19. The age- and sex-weighted excess mortality estimates are shown in **Figure 3**. The asymmetric distribution of excess deaths in 2020 and 2021 for Sweden vs. Denmark, Finland, and Norway is clear, as is the higher specific mortality for men, with the male mortality in Sweden during 2020 being particularly high. The uncertainties in the estimates are discussed further below.

**Table 3.** Estimated sex-and age-weighted excess deaths extrapolated from 2010−2019 death rates.

|  | Denmark | Finland | Iceland | Norway | Sweden |
| --- | --- | --- | --- | --- | --- |
| Men |  |  |  |  |  |
| Not age-weighted |  |  |  |  |  |
| 2020 | 681 | 659 | -47 | 594 | 4608 |
| 2021 | 1733 | 1654 | -60 | 1280 | 1852 |
| 2020+2021 | 2414 | 2313 | -107 | 1874 | 6460 |
| Age-weighted |  |  |  |  |  |
| 2020 | 666 | 657 | -24 | 249 | 4479 |
| 2021 | 1726 | 1632 | -40 | 776 | 1547 |
| 2020+2021 | 2392 | 2289 | -64 | 1025 | 6025 |
| Women |  |  |  |  |  |
| Not age-weighted |  |  |  |  |  |
| 2020 | 336 | 114 | -51 | 84 | 2895 |
| 2021 | 1727 | 905 | -61 | 1126 | -84 |
| 2020+2021 | 2062 | 1019 | -111 | 1211 | 2810 |
| Age-weighted |  |  |  |  |  |
| 2020 | 271 | 9 | -26 | -171 | 2667 |
| 2021 | 1614 | 767 | -32 | 710 | -573 |
| 2020+2021 | 1885 | 776 | -58 | 539 | 2095 |

**Figure 3.**
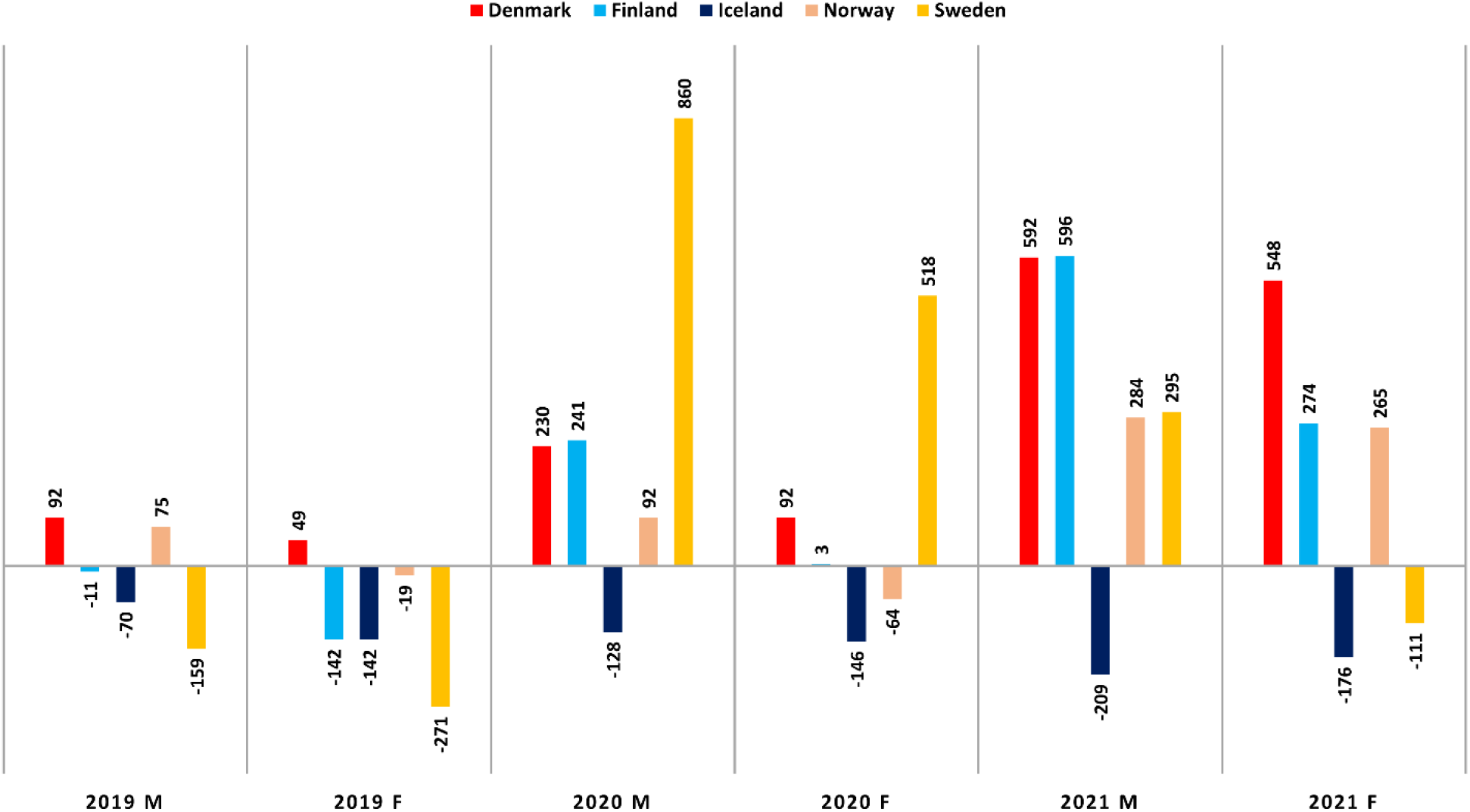
Estimated excess deaths per 1 million population, stratified by sex. The estimates for 2019 are also based on the trend of age- and sex-specific death rates for 2010−2019, scaled to 2019 mean populations of each age and sex group.

Since the periodic variations in the age sub-populations of women and men are largely synchronized (similar birth cycles), this is indicative of real mortality effects rather than a population size effect. We also find that age-correction effects (as e.g., affecting Norway and Denmark differently), when large, are seen for both sexes (**Table 3**), except Finland, where the correction mostly affects women.

### Comparison of old age mortalities

The age-standardized deaths in **Figure 2** enable direct comparison considering population structure differences but do not clarify whether the death rates are high or low relative to each country’s specific mortality expectations. This is important because mortality is determined by many drivers specific to each country, giving different age-specific death already before the pandemic (**Figure 1**).

**Figure 4** shows the age-specific mortality ratios based on age-adjusted deaths and the populations of each age group over time (actual death rate divided by the expected death rate in the age group). While age-specific mortality is steeply increasing with age and total excess deaths strongly dominated by 75+, the *relative* death rate changes during the pandemic were specific to different age groups in different countries. However, higher-than-expected death rates were seen in 2020 in Sweden among people of 80+ years and in 2021 in Denmark among people in the seventies. The Danish and Finnish mortalities were uniformly higher than expected in 2021 than in 2020 for all age groups.

**Figure 4.**
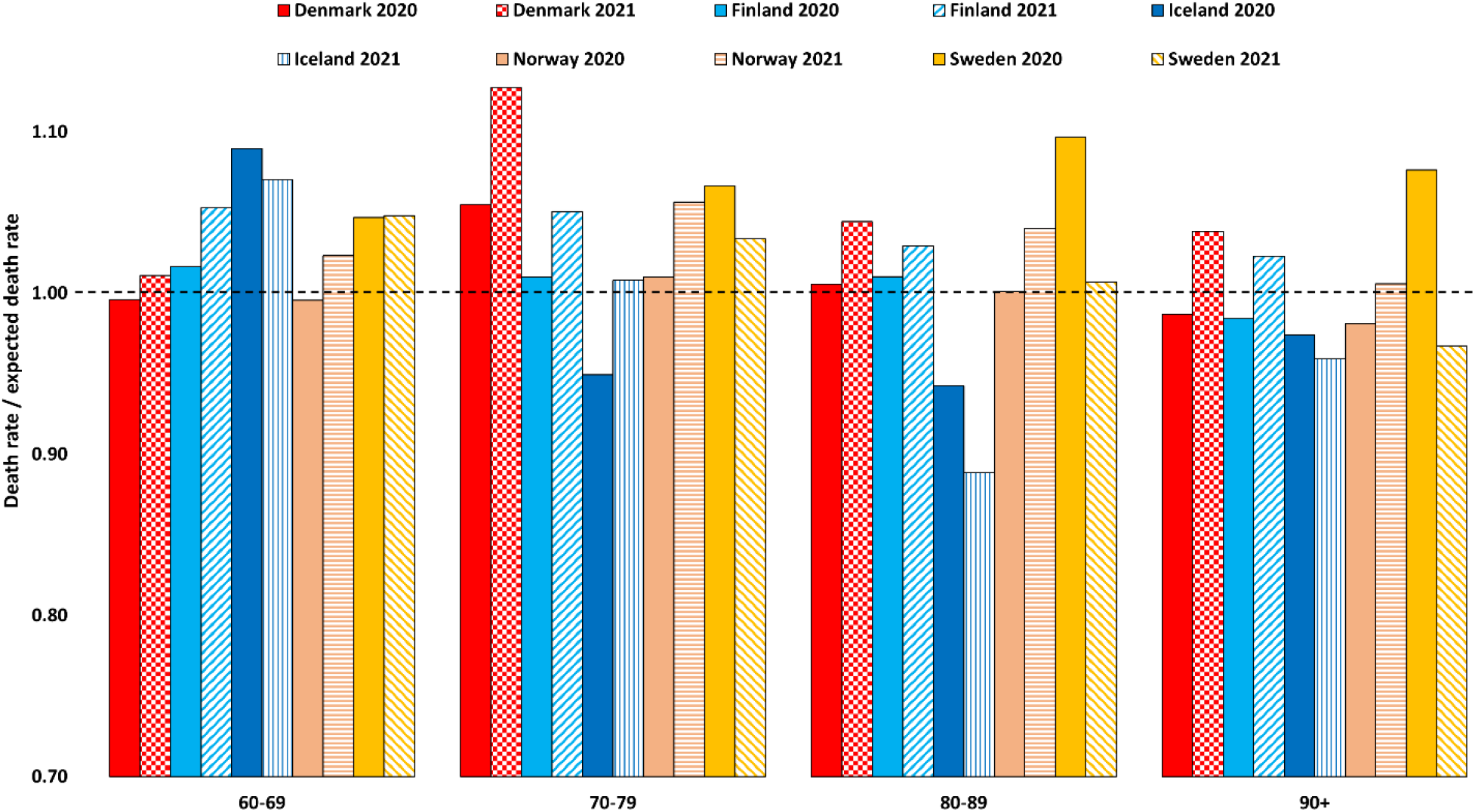
Age-specific death rates for 10-year age groups divided by expected age-specific death rates for the Nordic countries in 2020 and 2021 (from extrapolation from 2010-2019 to 2020 and 2021, taking the populations of the age groups of 2020 and 2021 into consideration).

The lower mortality in Sweden in 2021 relative to 2020 was largely driven by the age groups of 75 years and above. Since the method accounts reasonably for mortality displacement, this effect seems to be partly due to pandemic response. However, due to the reversed mortality patterns Sweden performed much better over the two-year period or in 2021 than if evaluated just for 2020. Selectively looking at one period could thus produce completely different interpretations and would neglect mortality displacement effects. Very low but also more uncertain mortalities were seen for Iceland in several age groups.

**Figure 5.** shows our estimated excess death rates (actual minus expected death rates) for all age groups, including 2018 and 2019 for comparison, using the same 2010−2019 linear model. These comparisons account for the increase in old age groups before and during the pandemic and document the inverse pattern for Sweden. Also, the excess death rate in Denmark in 2020 was substantially lower than the severe influenza year 2018, whereas 2018 and 2021 were similar in age-dependent impact when judged from this metric. The impact in Norway and Finland was also mainly visible in 2021. **Figure S7** summarizes the excess death estimates, excess death rates, and mortality ratios for all age groups.

**Figure 5.**
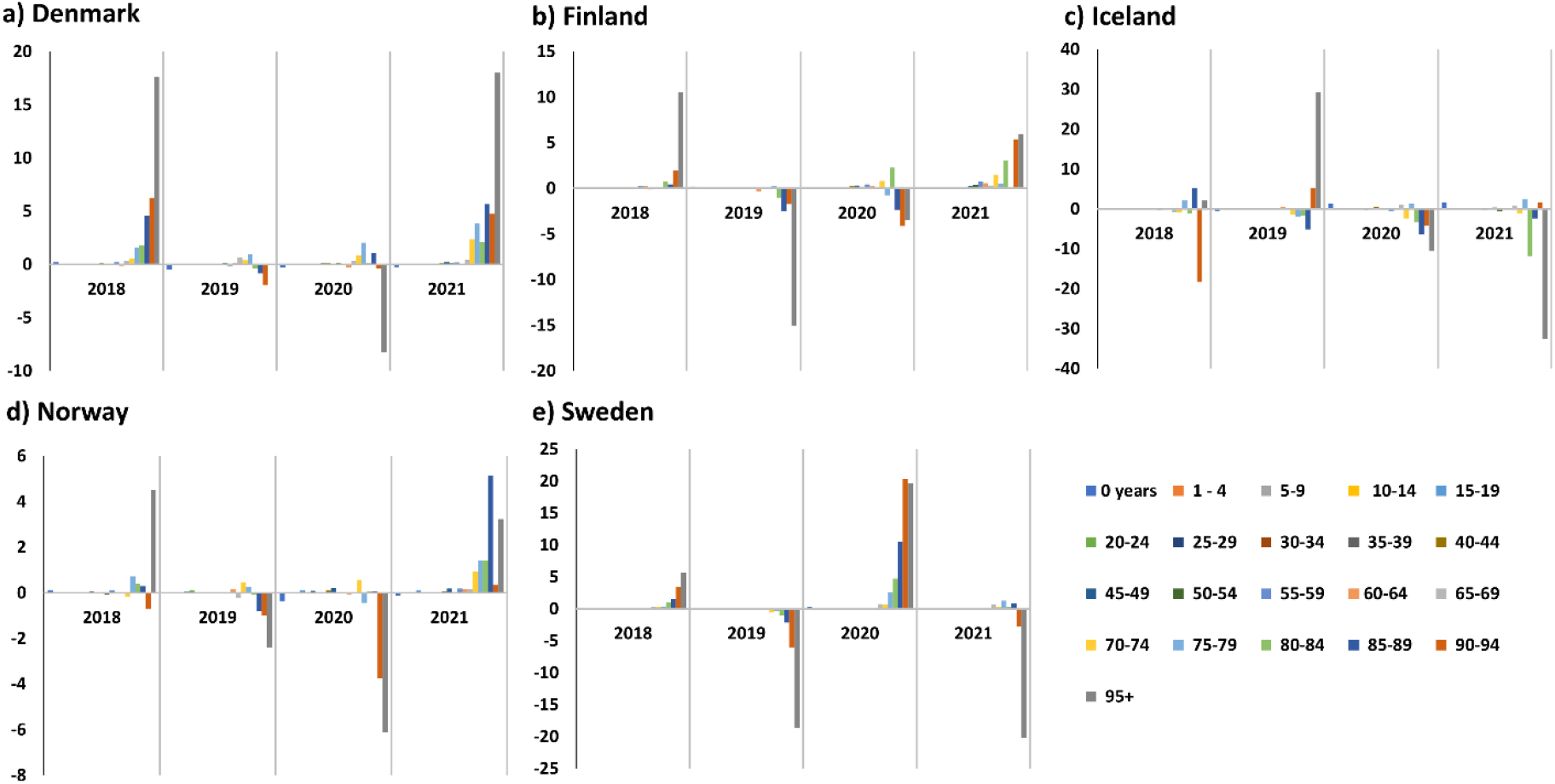
Age-specific excess death rates 2018, 2019, 2020, and 2021. **a)** Denmark. **b)** Finland. **c)** Iceland. **d)** Norway. **e)** Sweden.

### Infant mortality

**Figure 6.** shows the death rates of young age groups in the Nordic countries, excluding newborns (<1 year). Infant excess mortality ratios (actual death rates divided by the expected death rates) are summarized in **Figure S8** for boys (**Figure S8a**) and girls (**Figure 8b**; Iceland omitted due to very small numbers and large fluctuations). Despite these mortality ratios being substantially different from 1, and some excess implied for some 5−9-year groups and teenagers in 2021 (**Figure S8**), these come from death rate trends with fluctuations of the same magnitude as the effects (**Figure 6**). We therefore conclude that mortality patterns for children, including newborns, are consistent with pre-pandemic noise levels. Although some pediatric mortality impact cannot be excluded, we also doubt that it can be shown with the data, due to the fluctuations that reduce with more deaths at higher age (**Figure S7c, S7f, S7i, S7l, S7o**).

**Figure 6.**
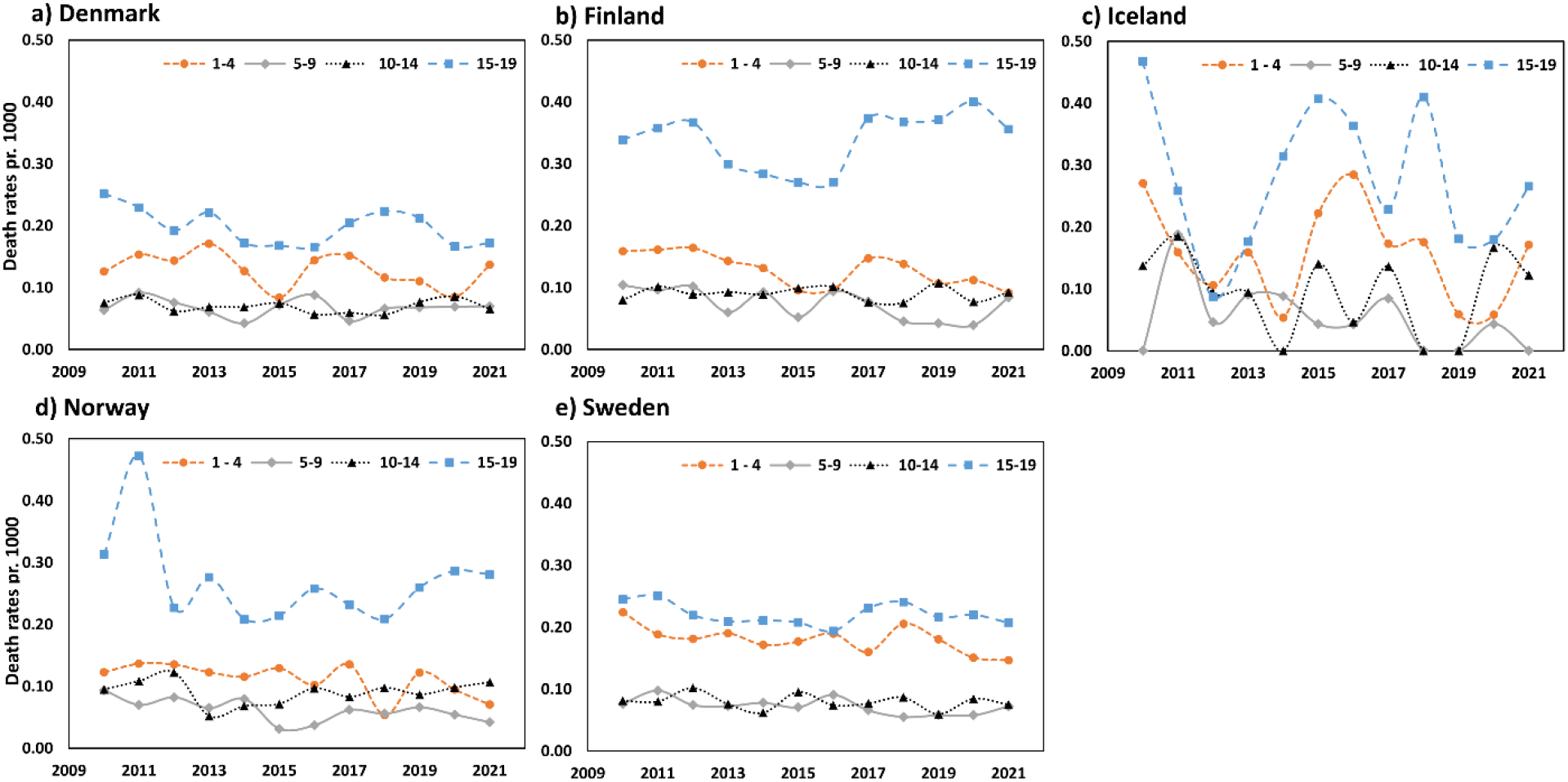
Death rates for children, 2010−2021. **a)** Denmark. **b)** Finland. **c)** Iceland. **d)** Norway. **e)** Sweden.

Specifically for newborns (age group <1 year), death rates are typically at the same level as approximately 45−54-year groups and 10−30 times higher than the average death rate of the next four years of life (1−4 years). However, this ratio differs substantially between the Nordic countries, with the highest death rates of newborns (defined as <1 year of age) seen in Denmark. For newborns, the death rates typically fluctuated around the pre-pandemic trends without a clear picture (**Figure S8**), with some mortality ratios for 2020 and 2021 above and below 1, whereas mortality ratios for 45−49y and 50−54y age groups were always above 1, except 50−54y for Iceland in 2021 (**Figure S9**). However, the fluctuations in the mid-age groups make these estimates more uncertain and contribute little to total excess death estimates (**Figure S10**).

## Discussion

### Main findings

We sought to examine the age and sex specific excess mortality in the Nordics during the pandemic. Our main findings suggest substantial mortality displacement, higher mortality for men than women, no significant mortality burden in children and young, and interesting country-specific variations in mortality patterns. Total age- and sex adjusted excess deaths, ratios of actual vs. expected death rates in country-specific mortality, and age-standardized mortality were all found to be useful metrics to assess pandemic mortality. Finland, Norway, and Denmark had most deaths in 2021 and Sweden in 2020. The death rates were particularly higher than expected among Swedes older than 80 year, and Danes older than 70 years. Our study also has several methodological implications, as discussed further below.

### Comparison to other models

We now compare our excess mortality estimates to other models and discuss sensitivities to reasonable variations in the data used. **Figure 7** compares our age- and sex-adjusted excess death estimates per million to estimates from the WHO excess mortality working group,^10^ IHME,^12^ the WMD model,^14^ the Economist model,^13^ the Bayesian model ensemble (BME),^6,32,33^ and the model by Levitt et al.^29^

**Figure 7.**
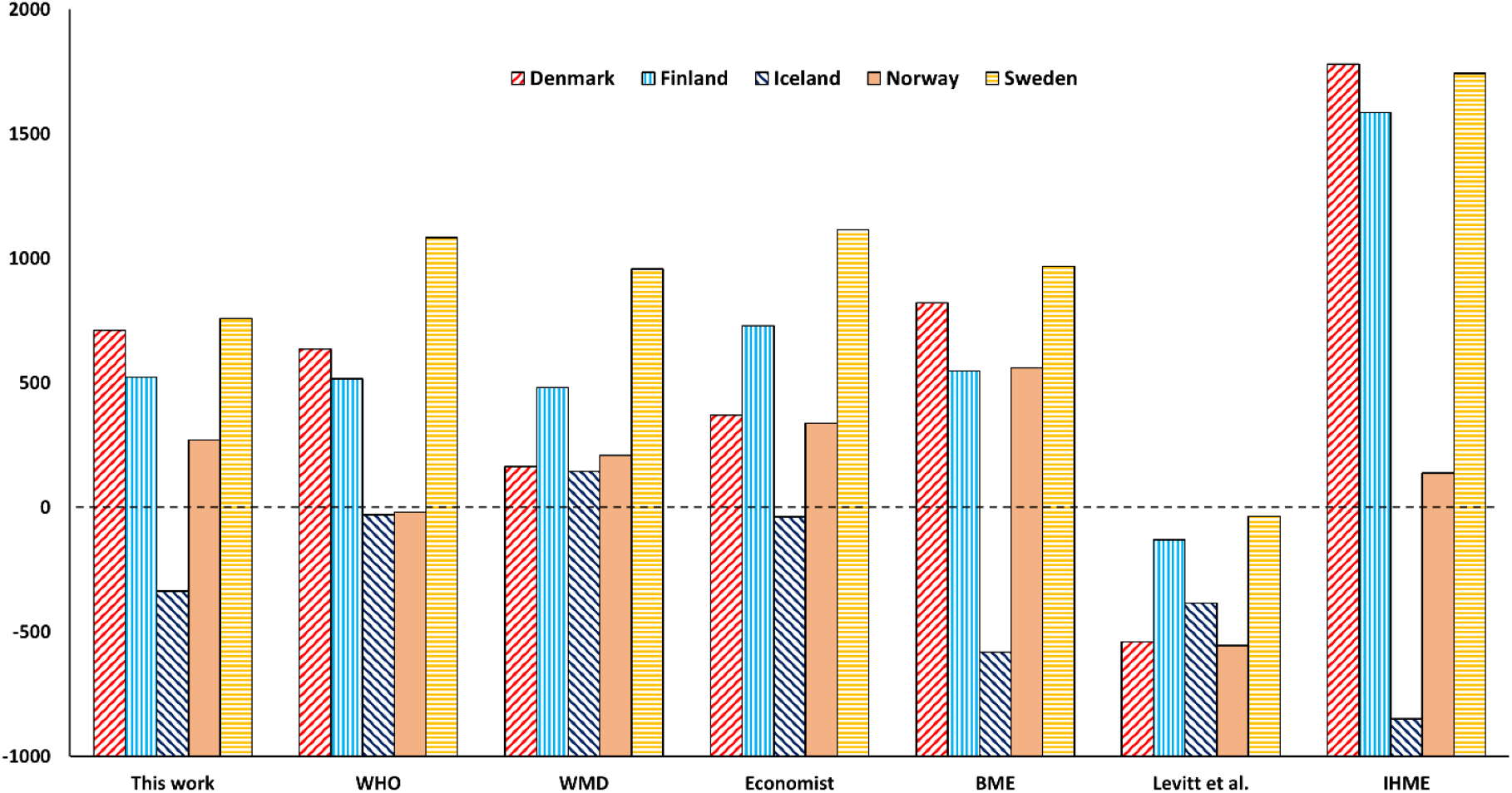
Comparison of excess death estimates for 2020 and 2021, per million population. All estimates divided by total mean populations of 2020 and 2021.

Our estimates are close to those of WHO^10,11^, especially for Denmark (4,277 vs. 3,716) and Finland (3,065 vs. 2,857). For Iceland, the difference of 111 people should be seen with the large uncertainty due to few deaths. The most notable differences are seen for Norway (1,564 this work; −101 from WHO) and somewhat for Sweden (8,120 this work; 11,255 WHO). Differences between deaths reported by the Nordic statistics departments and the WHO taskforce explain some of the discrepancy (**Table S3**). Still, the similarity is encouraging since our model is simple and parameter-free and only uses linear regression on actual death and population data. We note that a revision for Sweden has been reported remedying oversensitivity of WHO estimates to splines,^34^ but it is not clear how this affects the the four other countries.

Our estimates, despite correction for age and sex, are also relatively close to the estimates from the WMD^14^, Economist^13^ and BME models^32,33^, but quite distinct from the IHME and Levitt models. While we explained the discrepancy to the IHME model previously and believe this model has errors in estimating baselines that lead to substantial overestimation of Danish and Finnish excess deaths,^6^ the poor agreement with Levitt et al. is notable as this model used age-adjusted estimates as here, however based on fewer age groups and a shorter time-period of extrapolation.^29^ The reasons for these differences are analyzed below.

Our study can also be compared to life-expectation studies.^16,18^ The life expectancy deficit was calculated from the differences between observed life expectancy and a Lee–Carter mortality forecast using the death rates for the fourth quarter of 2015−2019^18^. These provide the best comparative mortality results so far but used relatively few age groups and 5-year data. While the metric differs, the two approaches agree on the historic mortality trend, mortality reversal from 2020 to 2021, little if any mortality effect in children, and Sweden experiencing comparatively less mortality in 2021 than 2020 compared to its neighbors.

For precise measures of excess mortality, we believe our estimates using fully commensurable data, no parameters, and a time-period that reduces mortality fluctuations of 2018 and 2019, but the conclusions are largely similar, which provides confidence to both approaches, and for perspective on more countries, without a need for very accurate excess death estimates for the Nordics, we thus recommend this work, which does not suffer from the horizontal baseline assumption (See below), as it allows for the trend in improving pre-pandemic mortality^18^.

An important observation from **Tables 1−3** is that the excess mortality is strongly asymmetric: Sweden experienced by far most excess mortality in 2020, whereas Norway, Finland, and Denmark experienced most in 2021. While this has been observed before and is visible also in the raw death series,^6^ these effects are robust to age- and sex stratification, supporting an effect of hybrid immunity, difference in vaccination roll-out, some remaining mortality displacement missed by the method, or other effects that warrant further study and more detailed data.

### Sensitivity tests and factors substantially influencing estimates

Below we discuss uncertainties in our estimates and relate our results to other models. We also refer to sensitivity tests performed in previous studies.^6,8^ Most notable is the impact of unusual mortality years before the crisis, which affect baselines obtained using different time periods. As an example, Denmark in 2018 had a severe influenza season, visible in **Figure 1**. We estimate 1,723 excess deaths in 2018 when using the 10-year age-specific method (i.e., interpolation for 2018, but extrapolation for 2020 and 2021), close to the estimate of 2017−2018 influenza deaths by the Danish Statens Seruminstitut (1,608).^35,36^

First, as an upper estimate of the impact of unusual mortality years and mortality displacement, we performed the same age-adjusted excess death estimates excluding 2019 (i.e., extrapolation from 2010−2018, **Table S4**). This led to a 12% higher excess death estimate for Denmark, 22% lower for Finland, 41% lower for Iceland, 12% higher for Norway, and 35% lower for Sweden. These estimates indicate plausible large impacts of mortality displacement and illustrate that a linear method, while having some of this included by averaging out fluctuations, does not account fully for it, and sensitivity tests are thus necessary. New models that account for mortality displacement seem warranted but are beyond the scope of our study.

Second, the impact of the population error (See methods) was estimated using January 1 start populations instead of mean populations of each age and sex group for 2020 and 2021. Since the subgroup populations change monotonic with irregularities reflecting birth cycles (**Figure S5**), simple linear extrapolation of expected subgroup populations is not possible, and it is arguably more accurate to use populations at the onset of the crisis years as a test (2020 and 2021). This effect was typically around 1% (from −0.8% to +1.2%) for the studied Nordic countries (**Table S5**) and thus not affecting the estimates much.

Third, while the Nordic countries had lower total excess deaths than many other countries^10,14,37^, the negative age-specific excess deaths for 2020−2021 by Levitt et al.^29^ are very distinct from other estimates (−3157 for Denmark, −716 for Finland, −142 for Iceland, −2994 for Norway, and −367 for Sweden).^29^ To understand the large discrepancy, we calculated the excess deaths using our 21 age groups (Levitt et al. used five age groups) and the data from the Nordic Council’s web page using the 3-year average in deaths 2017−2019 to estimate baselines and excess deaths (**Table S6**). The resulting age-specific excess deaths are close to those of Levitt et al. considering that we use somewhat different death and population data (−2541 for Denmark, −1507 for Finland, −196 for Iceland, −3110 for Norway, and 386 for Sweden).

We conclude that the negative excess deaths of Levitt et al.^29^ are erroneous and due to averaging the pre-pandemic baseline, when the age-specific death rates are declining. This leads to an overestimation of expected deaths and a substantial underestimation of excess deaths. A 5-year average, the time-period normally applied, would have made these estimates even more negative and implausible **(Table S7**), i.e., the choice of only three years for extrapolation by Levitt et al.^29^ partly reduces the error.

Fourth, many models somewhat arbitrarily use five years (2015−2019) to estimate baselines. The effect of using 5 instead of 10 years is seen in **Table S8** for excess deaths per million, comparing both 5-year average projection from death rates, 5-year linear projection from death rates, 5-year linear projection from raw deaths, 10-year projection, and the WMD estimates, which uses 5-year linear extrapolation of crude deaths to establish baselines.^14^ To ensure comparison, none of the estimates in **Table S8** are age-corrected.

As shown, despite minor differences due to different uses of death data, our 5-year trend reproduces quite well with those of the WMD model (**Table S8**).^14^ Use of death rates rather than crude deaths also has some effect. A 5-year trend puts very large emphasis on 2018 and 2019 mortalities, e.g., the high 2018 mortality in Denmark due to the severe influenza season,^38^ and this emphasis increases the baseline and reduced excess death estimates relative to a longer extrapolation. Such effects of time-period have been noted before.^8^ Since the trends are linear over the full extrapolation period a longer period is preferred, as 5-year trends are sensitive to 2018 and 2019.^6^ The same effect as in the WMD estimates on especially Denmark and Sweden was also seen by Demetriou et al. who also used 2015−2019 to estimate baselines.^39^

We have previously shown^6^ that IHME’s estimates for the Nordic countries^12^ are unreliable, with their expected deaths for 2020 and 2021 inconsistent with the pre-pandemic trends in the Nordic register data, in addition to other comparative anomalies.^6^ **Figure 2** puts IHME’s model in further age-specific context, with e.g., its 10,800 excess deaths for Denmark in 2020+2021,^12^ i.e., 1850 deaths per million or almost 1000/million per year, being neither consistent with non-age corrected data^6^ or age-corrected data. Thus while IHME substantially overestimates Nordic excess deaths,^6^ we also conclude that horizontal baseline methods are prone to major errors due to real trends in mortality, and e.g., Levitt et al.^29^ substantially underestimate excess deaths, with all other estimates clustering far from this and IHME (**Figure 7**). Pre-pandemic averaging is also used by EUROSTAT^40^ and similar methods^41,42^ and our findings may raise concerns about their accuracy too.

### Limitations and future work

The estimates provided here account for mortality displacement and outlier years better than other estimates published so far and additionally separate excess deaths into 42 age and sex groups, using final annualized register data without e.g. ISO calendar effects. Our work however still has several limitations. First of all, considerable uncertainty still relates to time period used^8^, but also, in terms of interpretation of excess deaths, other covariates relevant to interpretation of the outcomes than age and sex, such as socioeconomics or care status^43^. Since the sensitivities to time-period are larger than the confidence intervals of any specific model, reporting such intervals is not reflecting upon the true uncertainty in model estimates,^6^ and we have thus not reported these. Instead, our sensitivity analyses in the supplementary information serve as important indications of relevant uncertainty in our estimates.

While we believe the fluctuations in death rates for children and mid-age groups are too large to estimate excess mortalities accurately, we do see indications of excess mortality in mid-age groups that appear to differ between countries (**Table S9**). Our study does not analyze life years lost, only excess deaths for age groups in total, but the excess death distributions on age groups in **Table S9** suggest that Norway and Finland will become comparatively substantially worse than Denmark, Sweden, and Iceland in a life-year-lost analysis than in an excess death analysis, partly due to variations in birth cycles making age groups relatively different in size. Such studies are welcomed.

## Conclusions

We investigated the impact of sex and age on the pandemic excess mortality in the five Nordic countries, using final annualized register death data. Our estimates agree well with those by the WHO^44^ and life expectancy studies^18^, which is encouraging since our approach is conceptually simple, transparent, easily reproducible, and easily subject to sensitivity tests. Our study quantifies the mortality impact of the pandemic in the Nordic countries and indicates roles of mortality displacement, higher mortality for men, and important country-specific variations in mortality patterns. We show that three complementary metrics (total age- and sex adjusted excess deaths, ratios of actual vs. expected death rates in the context of each country’s mortality patterns, and age-standardized mortality) are all important to a total discussion of pandemic mortality outcome. Sweden had more excess deaths in 2020 than 2021 whereas Finland, Norway, and Denmark had the opposite. Higher-than-expected death rates were seen in 2020 in Sweden among 80+ and in 2021 in Denmark among people in the seventies. The pandemic put death rates back to 2015 for Sweden in 2020 but overall, death rates were still among the lowest ever in the Nordic countries during 2020 and 2021, consistent with a pre-pandemic trend of improving population health.

## Data Availability

All data required to reproduce this work are available at the web pages of Statistics Denmark, Statistics Norway, Statistics Sweden, Statistics Finland, and Statistics Iceland (Comparative Nordic data: mean population sizes, mortality rates).

https://pxweb.nordicstatistics.org/pxweb/en/Nordic%20Statistics/Nordic%20Statistics__Demography__Population%20change/

## Data availability

All data required to reproduce this work are available at the web pages of Statistics Denmark, Statistics Norway, Statistics Sweden, Statistics Finland, and Statistics Iceland, and public sites as summarized below:

Comparative Nordic data: mean population sizes, mortality rates: https://pxweb.nordicstatistics.org/pxweb/en/Nordic%20Statistics/Nordic%20Statistics__Demography__P__opulation%20change/

Populations January 1 divided on sex and age groups: https://pxweb.nordicstatistics.org/pxweb/en/Nordic%20Statistics/Nordic%20Statistics__Demography__P__opulation%20size/POPU01.px/

### Ethics approval

The study used anonymous, public total annual death and population data from the statistics departments of the Nordic countries, and thus did not require any ethics approval.

### Funding

No specific funding was received in relation to this work. J.B. was supported by grants for research on COVID-19 and pandemic preparedness from the Swedish Research Council (VR; grant number 2021–04665) and Sweden’s Innovation Agency (Vinnova; grant number AQ7 2021–02648).

## Supplementary information

**Table S1.** Comparison of deaths from Nordic Council (NC) using calendar years and WMD (World Mortality Dataset) using ISO weeks.

|  |  | 2015* | 2016 | 2017 | 2018 | 2019 | 2020* | 2021 |
| --- | --- | --- | --- | --- | --- | --- | --- | --- |
| **Denmark** | NC | 52555 | 52824 | 53261 | 55232 | 53958 | 54645 | 57152 |
|  | WMD | 53435 | 52581 | 53103 | 55076 | 53805 | 55478 | 57037 |
| **Finland** | NC | 52492 | 53923 | 53722 | 54527 | 53949 | 55488 | 57659 |
|  | WMD | 53467 | 53604 | 53516 | 54369 | 53820 | 56237 | 57580 |
| **Iceland** | NC | 2178 | 2309 | 2240 | 2257 | 2277 | 2307 | 2333 |
|  | WMD | 2223 | 2295 | 2229 | 2247 | 2261 | 2344 | 2333 |
| **Norway** | NC | 40727 | 40726 | 40774 | 40840 | 40684 | 40611 | 42002 |
|  | WMD | 41528 | 40436 | 40631 | 40704 | 40458 | 41190 | 41881 |
| **Sweden** | NC | 90907 | 90982 | 91972 | 92185 | 88766 | 98124 | 91958 |
|  | WMD | 92555 | 90502 | 91627 | 91920 | 88545 | 99654 | 91509 |

\* ISO leap years. NC = Official annual death data using calendar years from the Nordic Council.
https://pxweb.nordicstatistics.org/pxweb/en/Nordic%20Statistics/Nordic%20Statistics_Demography_Population%20change/DEAT01.px/. WMD = deaths assigned to each year by the World Mortality Dataset. https://github.com/akarlinsky/world_mortality/blob/main/world_mortality.csv

**Table S2.** Standard populations.

| Age group | Segi standard | Scandinavian standard | WHO Standard | Denmark 2020 |
| --- | --- | --- | --- | --- |
| 0–4 | 12 | 8 | 8.86 | 5.30 |
| 5–9 | 10 | 7 | 8.69 | 5.20 |
| 10–14 | 9 | 7 | 8.6 | 5.81 |
| 15–19 | 9 | 7 | 8.47 | 5.87 |
| 20–24 | 8 | 7 | 8.22 | 6.50 |
| 25–29 | 8 | 7 | 7.93 | 6.89 |
| 30–34 | 6 | 7 | 7.61 | 6.23 |
| 35–39 | 6 | 7 | 7.15 | 5.63 |
| 40–44 | 6 | 7 | 6.59 | 6.16 |
| 45–49 | 6 | 7 | 6.04 | 6.75 |
| 50–54 | 5 | 7 | 5.37 | 6.98 |
| 55–59 | 4 | 6 | 4.55 | 6.76 |
| 60–64 | 4 | 5 | 3.72 | 5.94 |
| 65–69 | 3 | 4 | 2.96 | 5.48 |
| 70–74 | 2 | 3 | 2.21 | 5.51 |
| 75–79 | 1 | 2 | 1.52 | 4.26 |
| 80–84 | 0.5 | 1 | 0.91 | 2.60 |
| 85–89 | 0.5 | 0.69* | 0.44 | 1.38 |
| 90–94 |  | 0.24* | 0.15 | 0.60 |
| 95+ |  | 0.07* | 0.045 | 0.18 |
| **Total** | 100 | 100 | 100 | 100 |

\* Original value of 1 for 85+ distributed and normalized based on WHO population.

**Table S3.** Total deaths tabulated by WHO excess mortality team vs. the Nordic statistics agencies (NC).

|  |  | WHO | NC | difference % |
| --- | --- | --- | --- | --- |
| Denmark | 2020 | 54790 | 54645 | 0.26 |
|  | 2021 | 57363 | 57152 | 0.37 |
| Finland | 2020 | 56927 | 55488 | 2.53 |
|  | 2021 | 58944 | 57659 | 2.18 |
| Iceland | 2020 | 2382 | 2307 | 3.15 |
|  | 2021 | 2413 | 2333 | 3.32 |
| Norway | 2020 | 40866 | 40611 | 0.62 |
|  | 2021 | 42046 | 42002 | 0.10 |
| Sweden | 2020 | 102022 | 98124 | 3.82 |
|  | 2021 | 96755 | 91958 | 4.96 |

**Table S4.**
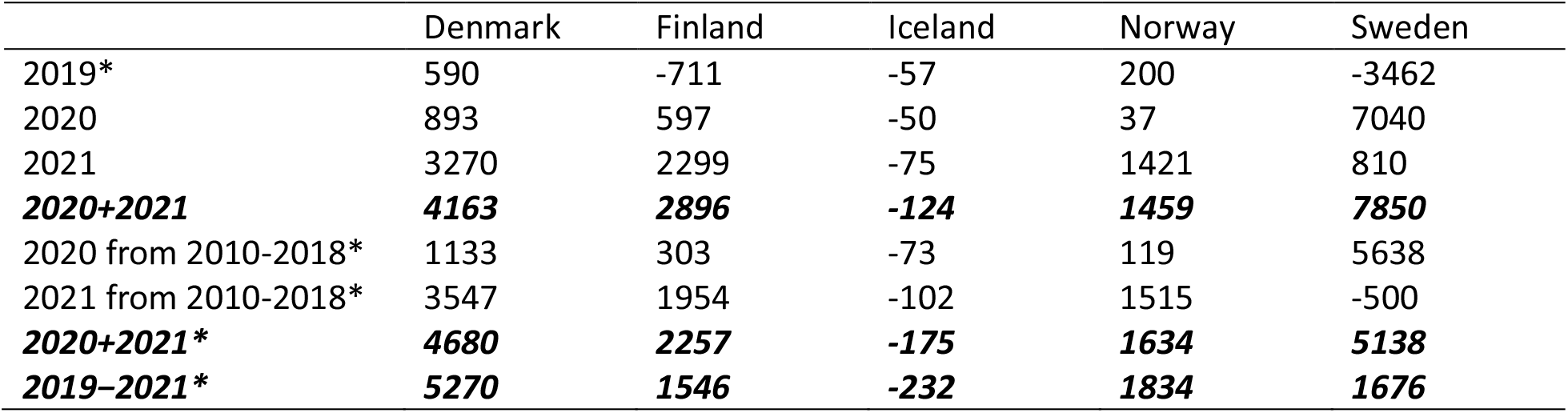
Impact on age-specific excess deaths of using 2010-2018 (*) instead of 2010-2019 for extrapolating expected deaths.

|  | Denmark | Finland | Iceland | Norway | Sweden |
| --- | --- | --- | --- | --- | --- |
| 2019* | 590 | -711 | -57 | 200 | -3462 |
| 2020 | 893 | 597 | -50 | 37 | 7040 |
| 2021 | 3270 | 2299 | -75 | 1421 | 810 |
| **2020+2021** | **4163** | **2896** | **-124** | **1459** | **7850** |
| 2020 from 2010-2018* | 1133 | 303 | -73 | 119 | 5638 |
| 2021 from 2010-2018* | 3547 | 1954 | -102 | 1515 | -500 |
| **2020+2021\*** | **4680** | **2257** | **-175** | **1634** | **5138** |
| **2019-2021\*** | **5270** | **1546** | **-232** | **1834** | **1676** |

**Table S5.** Comparison of using start vs. mean populations of 2020 and 2021.

|  | Denmark | Finland | Iceland | Norway | Sweden |
| --- | --- | --- | --- | --- | --- |
| Age-specific |  |  |  |  |  |
| 2020 mean pop | 893 | 597 | -50 | 37 | 7040 |
| 2021 mean pop | 3270 | 2299 | -75 | 1421 | 810 |
| **2020+2021 mean pop** | **4163** | **2896** | **-124** | **1459** | **7850** |
| 2020 start pop | 881 | 611 | -50 | 42 | 6993 |
| 2021 start pop | 3234 | 2279 | -73 | 1406 | 800 |
| **2020+2021, start pop** | **4115** | **2890** | **-123** | **1448** | **7792** |
| Age/sex-specific |  |  |  |  |  |
| 2020 mean pop | 937 | 666 | -50 | 79 | 7146 |
| 2021 mean pop | 3340 | 2399 | -72 | 1485 | 974 |
| **2020+2021, mean pop** | **4277** | **3065** | **-122** | **1564** | **8120** |
| 2020 start pop | 923 | 681 | -49 | 83 | 7093 |
| 2021 start pop | 3302 | 2374 | -71 | 1470 | 959 |
| **2020+2021, start pop** | **4226** | **3055** | **-121** | **1553** | **8052** |

**Table S6.** Total estimated excess deaths from expected deaths in 2020+2021 interpolated from 3-year average in death rates, scaled to known mean populations of 2020 and 2021. Because of the declining mortality rates, this method overestimates expected deaths and underestimates excess deaths.

|  | Denmark | Finland | Iceland | Norway | Sweden |
| --- | --- | --- | --- | --- | --- |
| Not age-specific |  |  |  |  |  |
| 2020 | 118 | 1281 | -43 | -672 | 5495 |
| 2021 | 2388 | 3339 | -56 | 497 | -1229 |
| Total | 2505 | 4620 | -99 | -175 | 4267 |
| Age-specific |  |  |  |  |  |
| 2020 | -1880 | -1149 | -74 | -1790 | 4051 |
| 2021 | -661 | -358 | -122 | -1319 | -3665 |
| Total | -2541 | -1507 | -196 | -3110 | 386 |
| Levitt et al.<sup>29</sup> | -3157 | -716 | -142 | -2994 | -367 |

**Table S7.** Total estimated excess deaths from expected deaths in 2020+2021 interpolated from 5-year average in death rates, scaled to known mean populations of 2020 and 2021.

|  | Denmark | Finland | Iceland | Norway | Sweden |
| --- | --- | --- | --- | --- | --- |
| Not age-specific |  |  |  |  |  |
| 2020 | 388 | 1518 | -90 | -970 | 4351 |
| 2021 | 2660 | 3577 | -104 | 198 | -2379 |
| Total | 3048 | 5094 | -194 | -772 | 1972 |
| Age-specific |  |  |  |  |  |
| 2020 | -2421 | -2071 | -132 | -2503 | 2584 |
| 2021 | -1215 | -1301 | -181 | -2047 | -5152 |
| Total | -3635 | -3372 | -313 | -4550 | -2567 |

**Table S8.** Estimated excess deaths per million, using 5-year and 10-year linear trends in death rates, scaled to mean populations of 2020 and 2021, or average of 2015−2019 without age-correction.

|  | Denmark | Finland | Iceland | Norway | Sweden |
| --- | --- | --- | --- | --- | --- |
| 5-year avg., death rates | 522 | 920 | -525 | -143 | 190 |
| 5-year trend, death rates | 257 | 601 | 192 | 255 | 1166 |
| **5-year trend, raw deaths** | **174** | **585** | **89** | **203** | **993** |
| **WMD<sup>14</sup>** | **165** | **481** | **143** | **209** | **957** |
| 10-year trend, death rates | 765 | 601 | -594 | 572 | 895 |

**Table S9.**
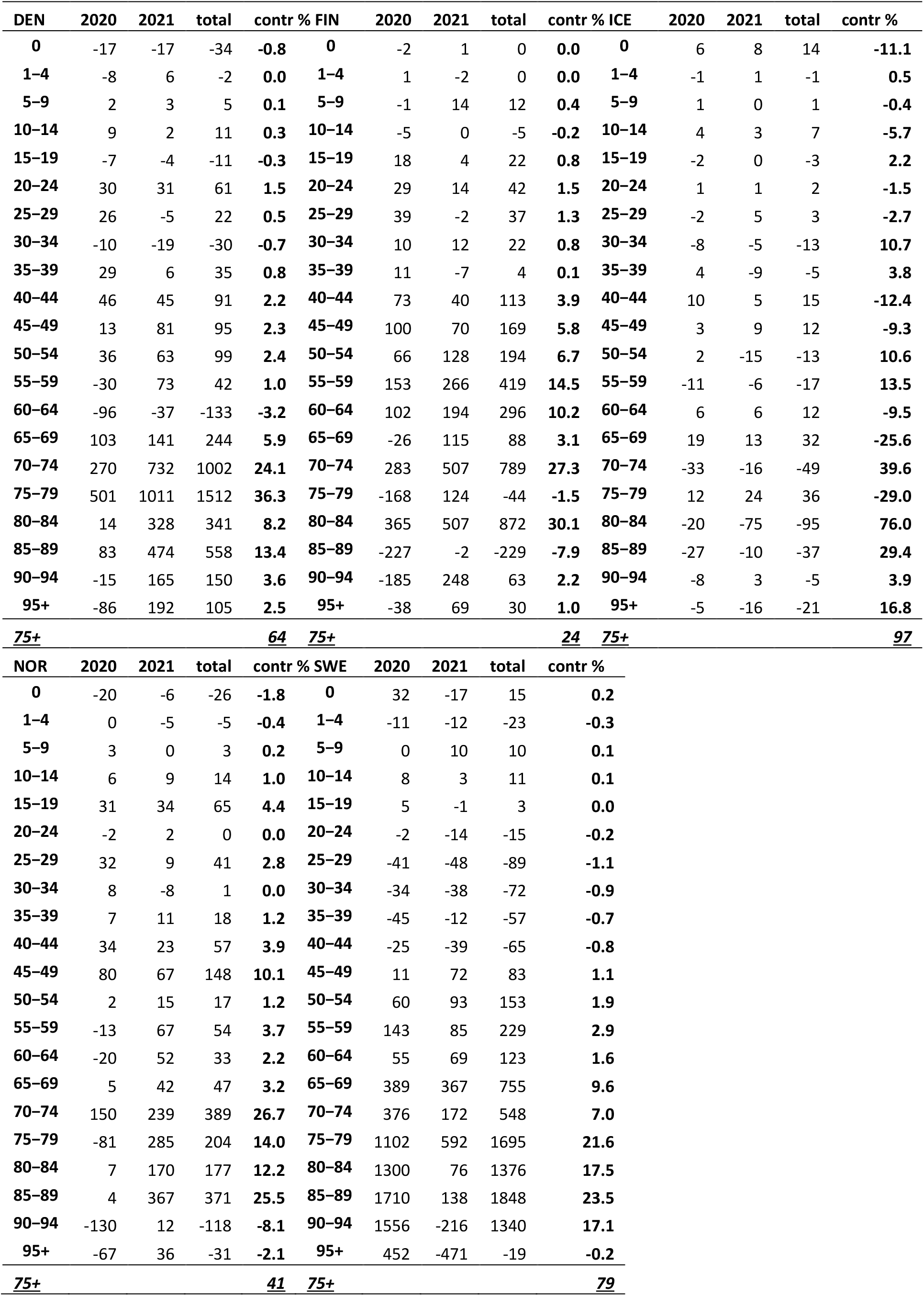
Estimated excess deaths distributed on age groups.

**Figure S1.**
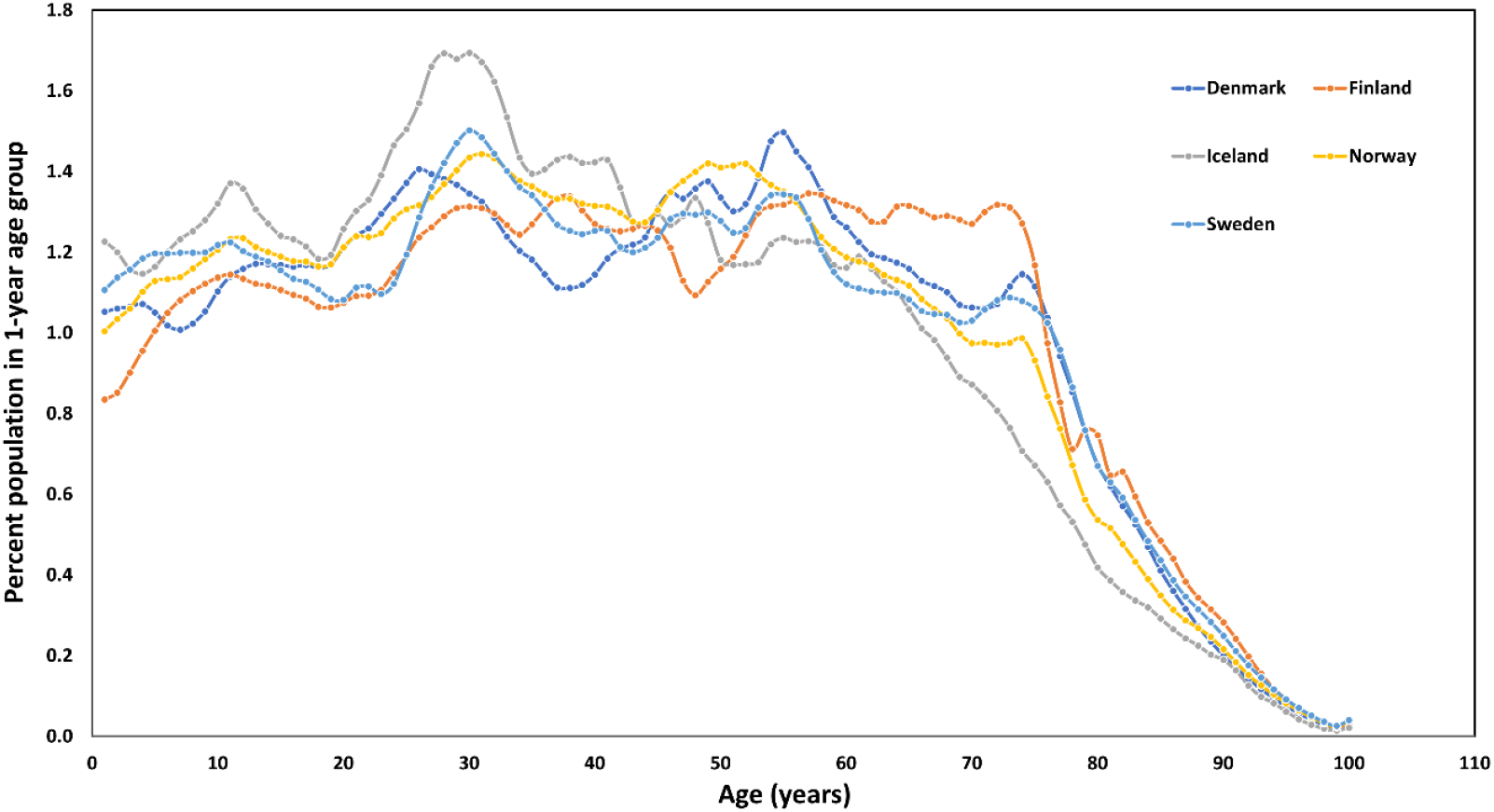
Mean population size per 1-year age group for the year 2020 when the pandemic started.

**Figure S2.**
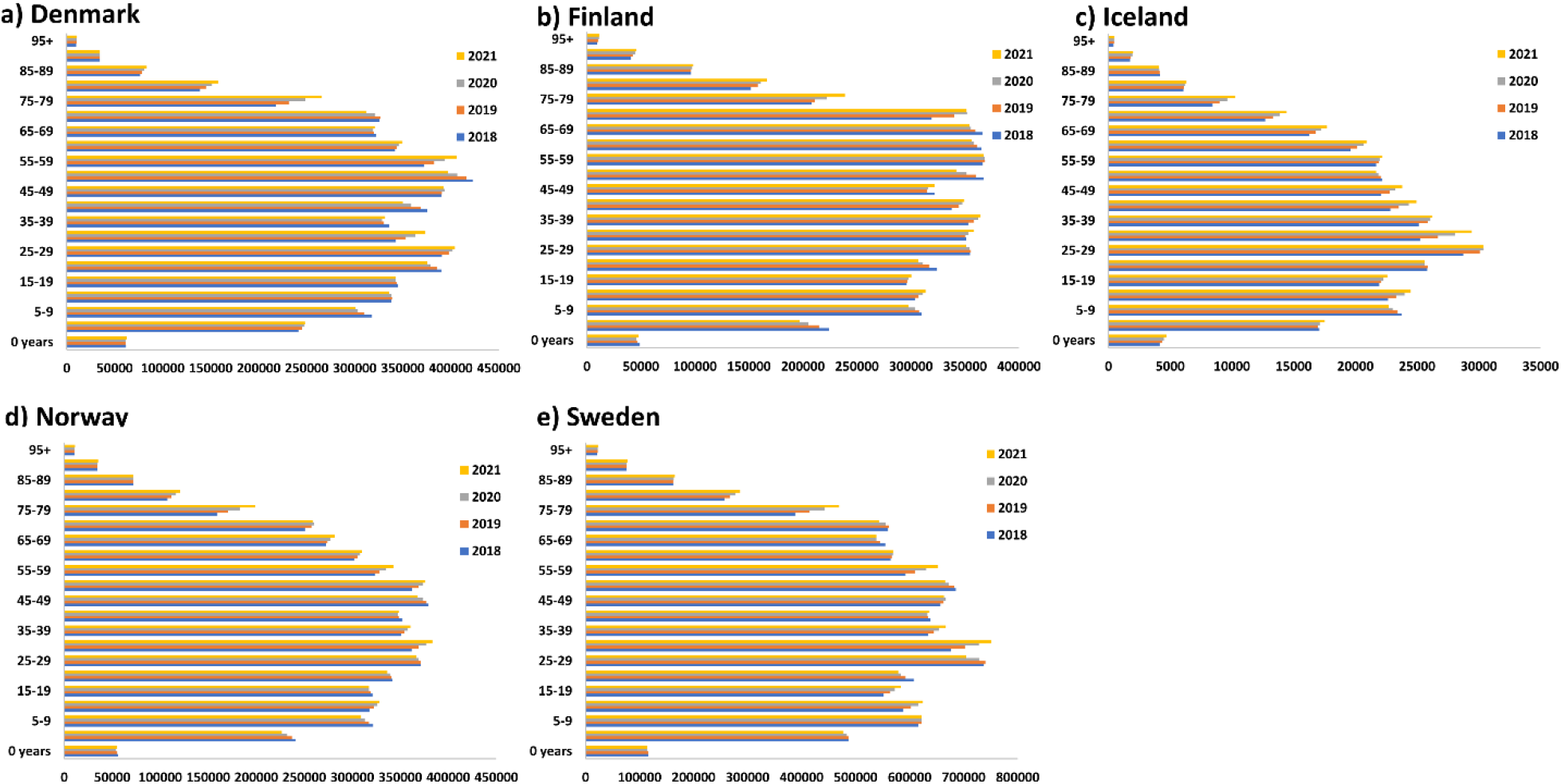
Mean populations in 2018, 2019, 2020, and 2021 per age group for Nordic countries. **a)** Denmark. **b)** Finland. **c)** Iceland. **d)** Norway. **e)** Sweden. Especially the size of the 75-79-year subgroup, which contributes highly to total excess deaths, has expanded over recent years due to a similar demographic change, related to Nordic birth cycles.

**Figure S3.**
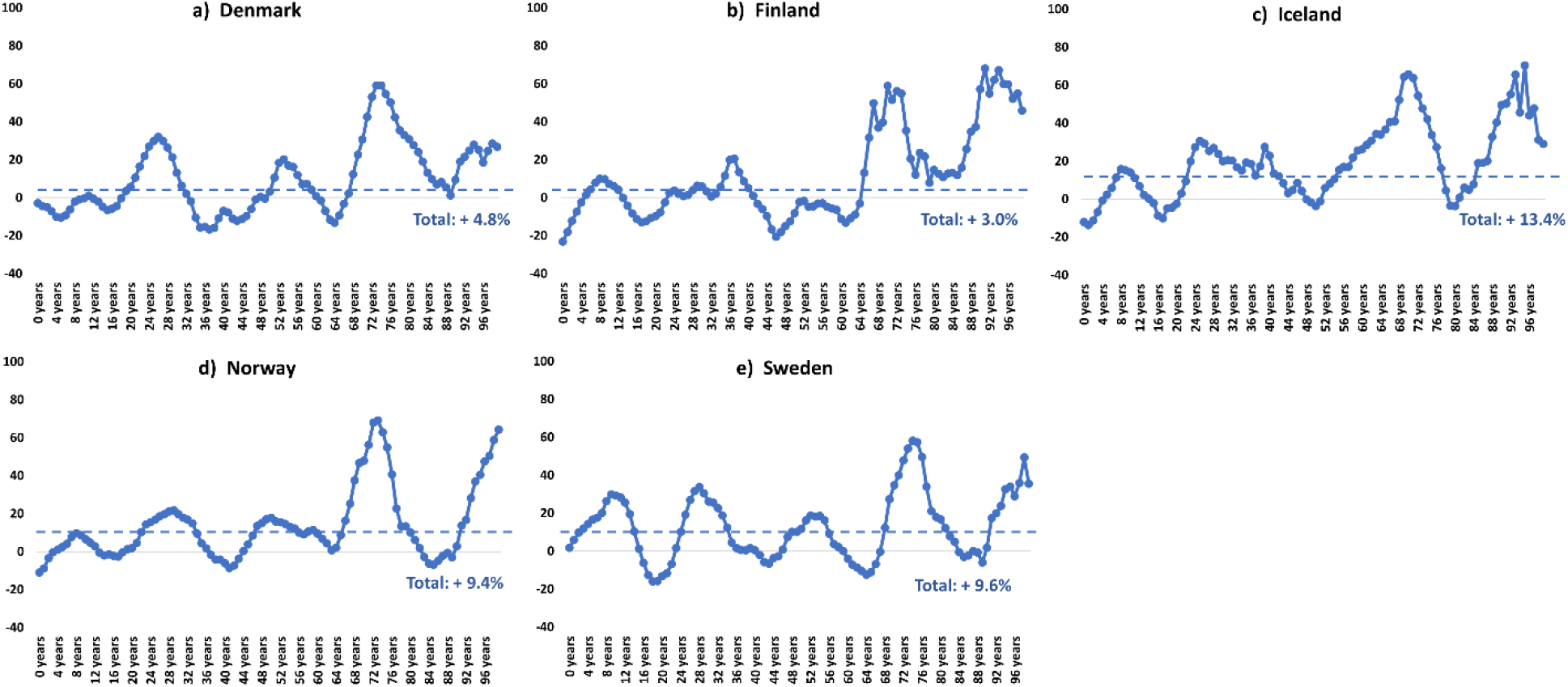
Impact of birth cycles of the Nordic countries on changing the size of age groups over time. Changes in population (in %) of 1-year age groups for the Nordic countries 2010−2019. **a)** Denmark. **b)** Finland. **c)** Iceland. **d)** Norway. **e)** Sweden. The stipulated blue lines show total population change. Source: https://pxweb.nordicstatistics.org/pxweb/en/Nordic%20Statistics/Nordic%20Statistics__Demography__Population%20size/POPU02.px/

**Figure S4.**
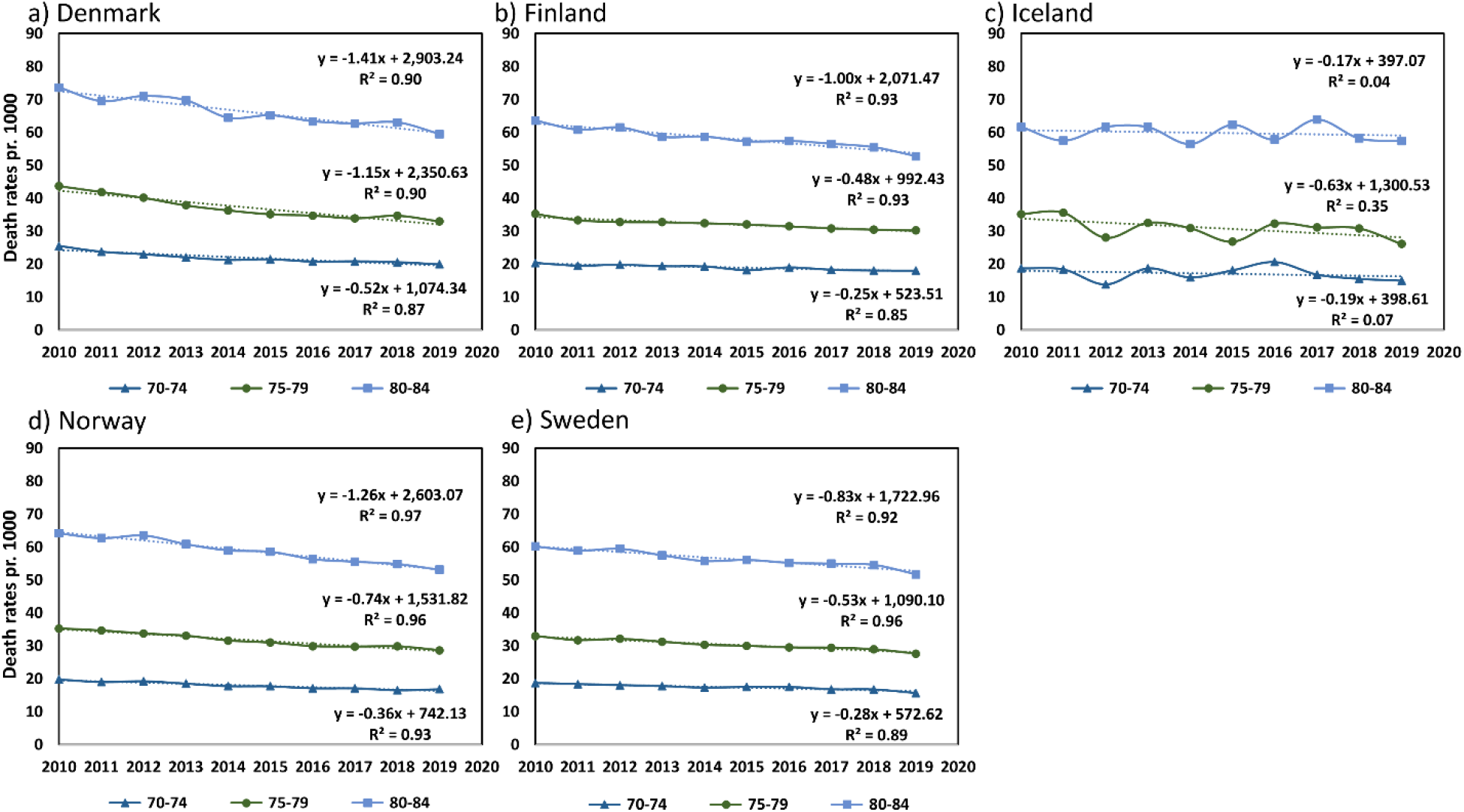
Linear trend lines in death rates for 70−74y, 75−79y, and 80-84y for 2010−2019. The linear death rates for Iceland are substantially fluctuating and have only small declines whereas Denmark, Finland, Norway, and Sweden have declines with linear fits showing R^2^ = 0.85−0.97. **a)** Denmark. **b)** Finland. **c)** Iceland. **d)** Norway. **e)** Sweden.

**Figure S5.**
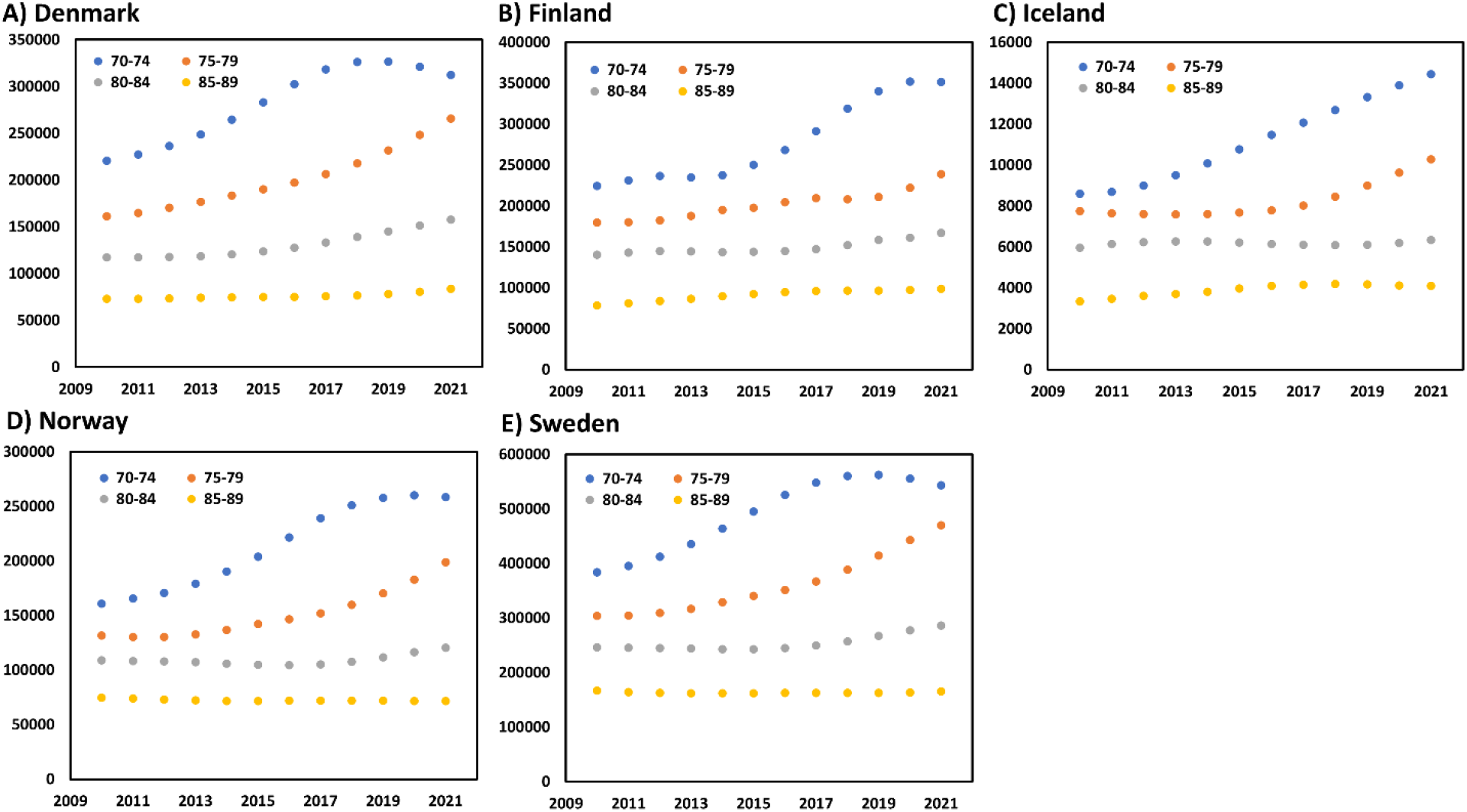
Populations of 5-year groups contributing most to total excess deaths, 2010−2021. **a)** Denmark. **b)** Finland. **c)** Iceland. **d)** Norway. **e)** Sweden.

**Figure S6.**
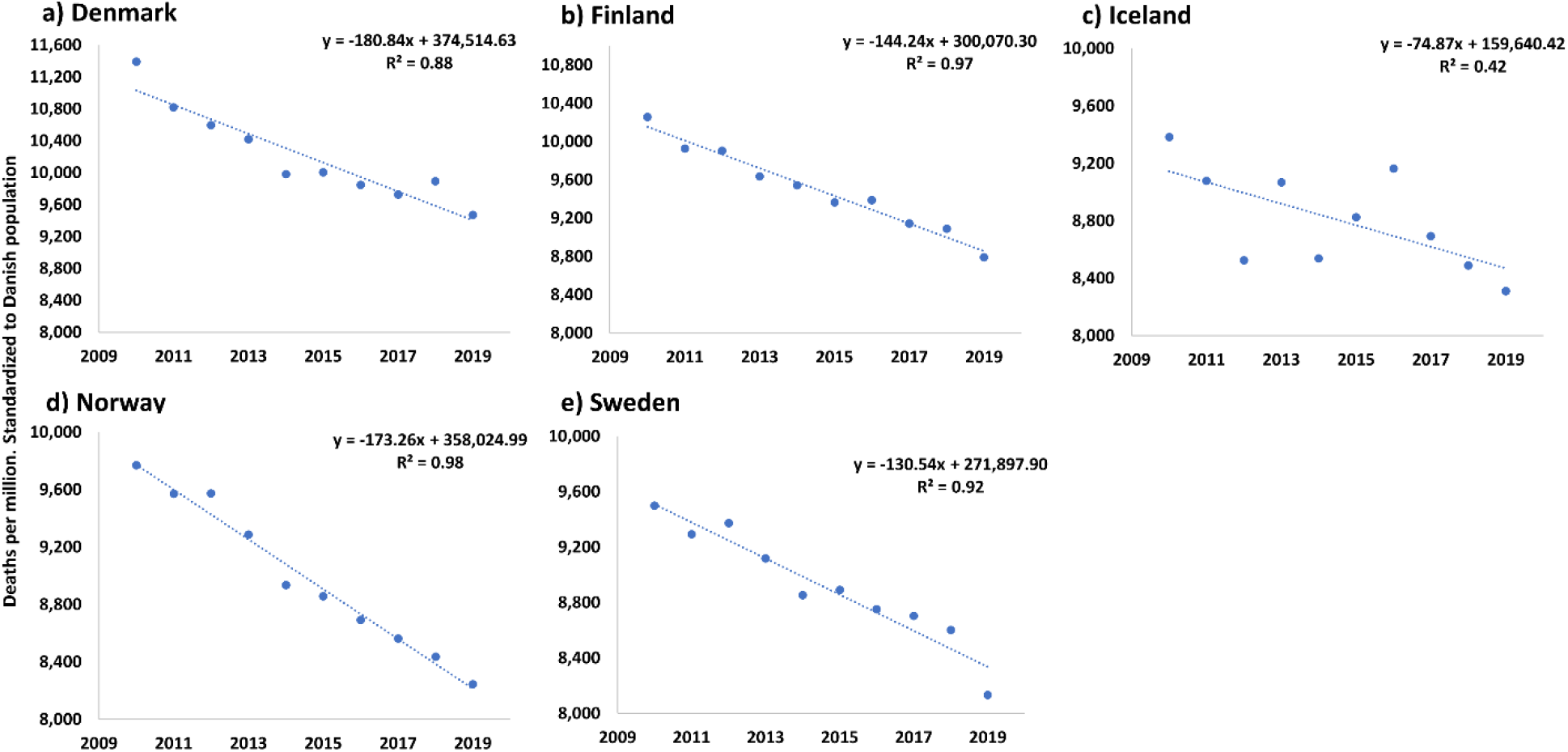
Trend in age-standardized age-specific death rates standardized to the Danish 2020 population, 2010−2021. **a)** Denmark. **b)** Finland. **c)** Iceland. **d)** Norway. **e)** Sweden.

**Figure S7.**
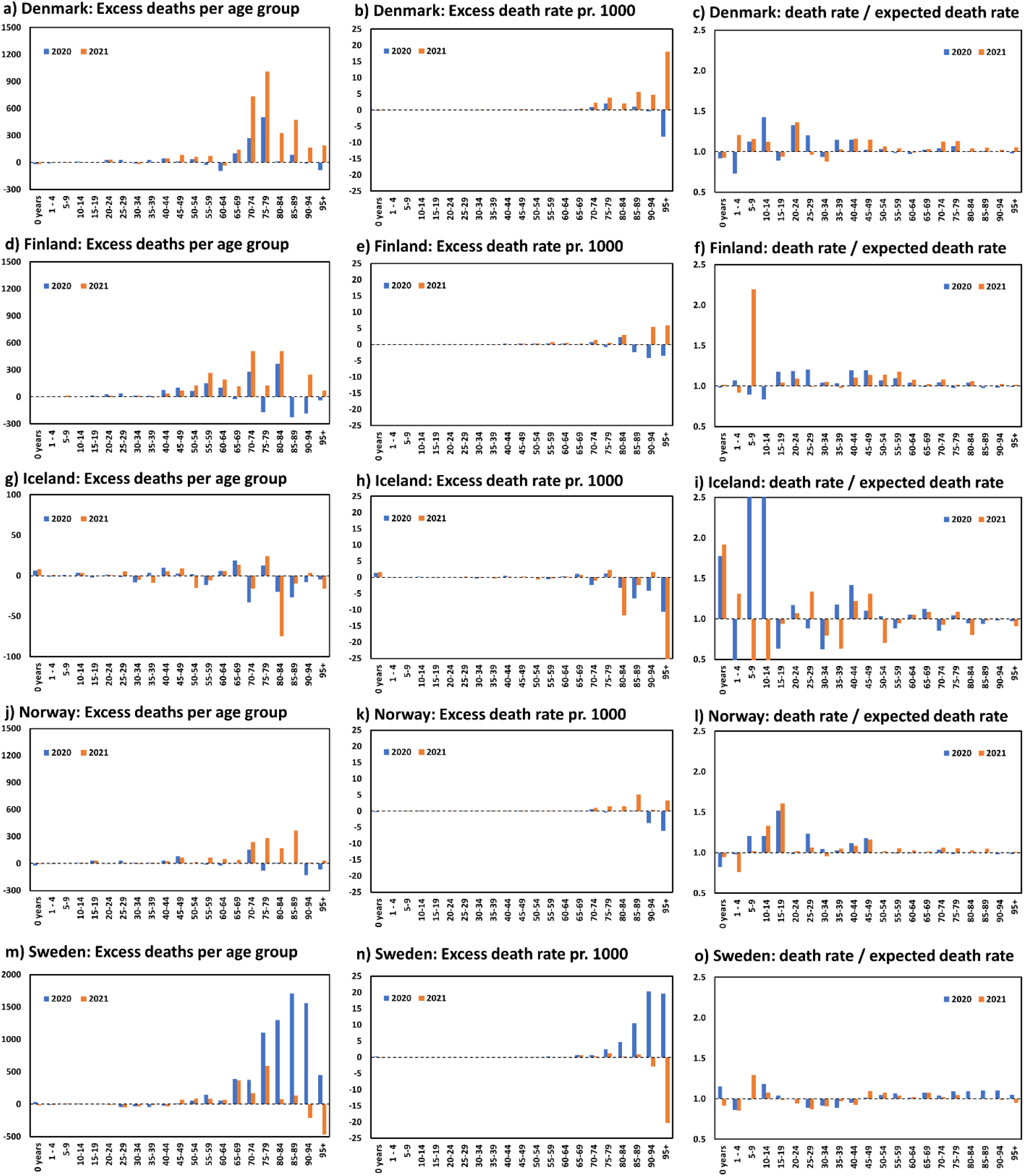
Excess death estimates (left), excess death rates (middle), and mortality ratios (death rates / expected death rates, right) for all age groups of the five Nordic countries. **(a-c)** Denmark. **(b-f**) Finland. **(g-i)** Iceland. **(j-l)** Norway. **(m-o)** Sweden.

**Figure S8.**
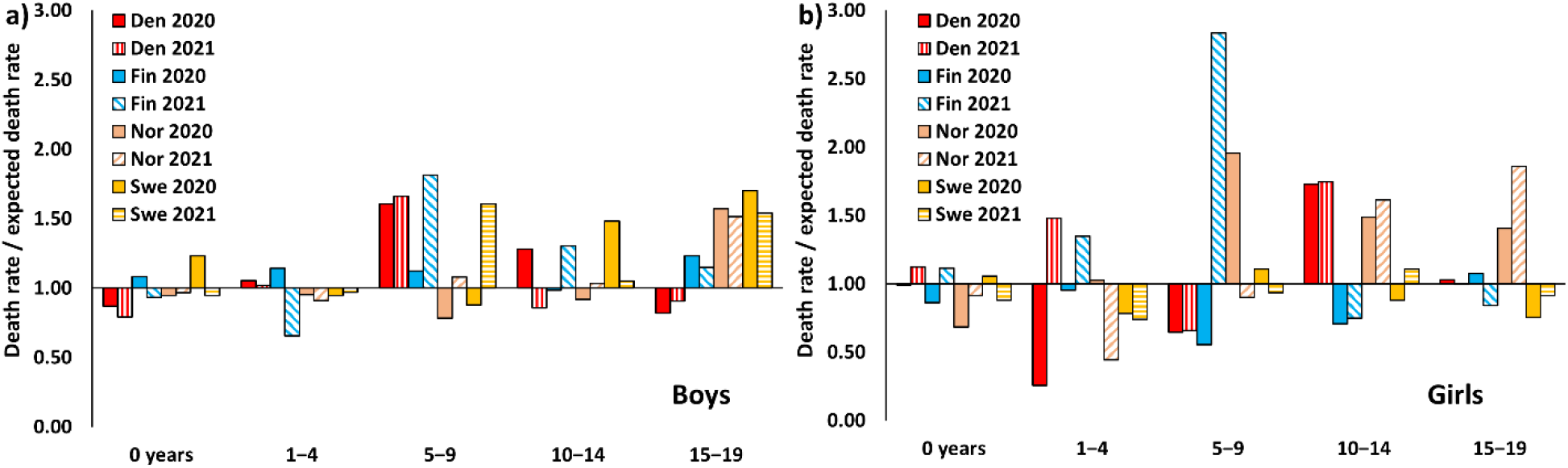
Actual death rates divided by expected death rates for children, 2020−2021. **A)** Boys. **B)** Girls. (Iceland omitted due to very large fluctuations and small numbers, including zero).

**Figure S9.**
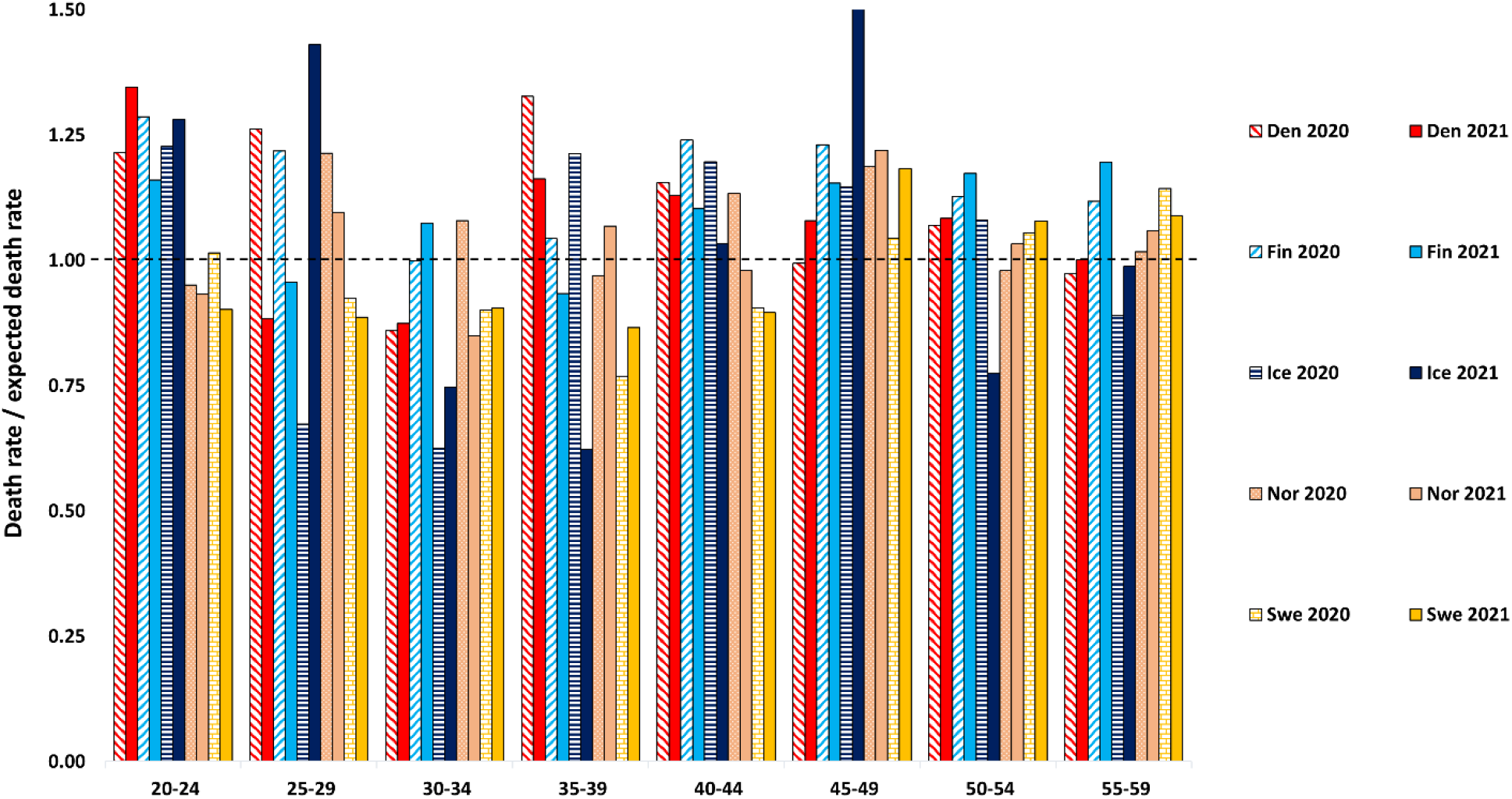
Actual death rates divided by expected death rates for 20−59-year groups, 2020−2021.

**Figure S10.**
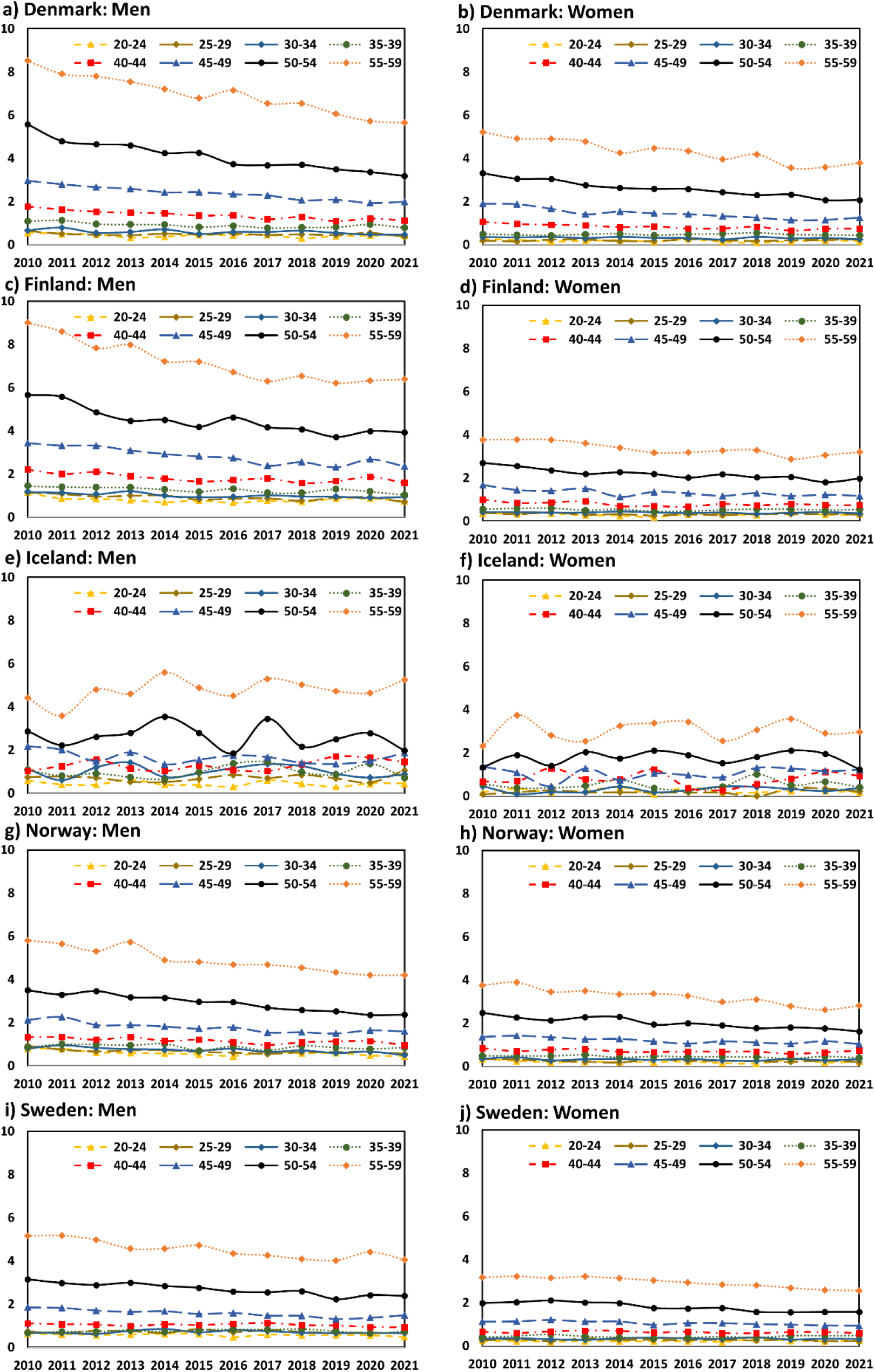
Observed death rates 2020−2021 for the Nordic countries. **a)** Denmark, men. **b)** Denmark, women. **c)** Finland, men. **d)** Finland, women. **e)** Iceland, men. **f)** Iceland, women. **g)** Norway, men. **h)** Norway, women. **i)** Sweden, men. **j)** Sweden, women.

